# Diagnostic Test Accuracy of Commercially Available Tests for The Recurrence of Bladder Cancer: A Systematic Review and Meta-Analysis

**DOI:** 10.64898/2026.02.02.26344871

**Authors:** Evangelia Ntzani, Konstantina-Eleni Tsarapatsani, Georgios-Alexandros Asimakopoulos, Hawre Jalal, Stella K. Kang, Tom Trikalinos, CISNET Bladder Cancer Modelling Investigators

**Affiliations:** Department of Hygiene and Epidemiology, University of Ioannina School of Medicine, Greece; Center for Evidence Synthesis in Health, Brown University School of Public Health, RI, USA; School of Epidemiology and Public Health, University of Ottawa, Ottawa, ON, Canada; Department of Radiology, Columbia University Irving Medical Center, New York, NY, USA

**Keywords:** Bladder cancer, urinary biomarker tests, recurrence, surveillance, meta-analysis

## Abstract

**Objective:** Bladder cancer (BC) is the most common malignancy of the urinary system and among the most frequently diagnosed cancers worldwide. This systematic review and meta-analysis aimed to evaluate the diagnostic accuracy of commercially available urinary biomarkers tests (UBTs) for detecting BC recurrence, focusing on pooled sensitivity and specificity estimates across different tests.

**Methods:** A systematic search was performed on PubMed and EMBASE up to May 2025 to identify studies assessing recurrence of BC in previously diagnosed patients using non-FDA approved UBTs, including Xpert Bladder Cancer, Bladder Epicheck, ADXbladder and Uromonitor. Eligible studies were synthesized using the bivariate Generalized Linear Mixed Model (GLMM) model.

**Results:** Out of 307 initially screened citations, 33 studies met the eligibility criteria, encompassing a total of 10,478 patients. Xpert Bladder Cancer was evaluated on 13 studies and Bladder Epicheck was assessed on 10 studies. ADXbladder and Uromonitor were assessed in four and six studies, respectively. Meta-analyses included 13 studies for Xpert Bladder Cancer and 10 studies for Bladder Epicheck, yielding pooled sensitivity (95% CI) and specificity (95% CI) estimates of 0.71 (0.61-0.79) and 0.78 (0.74-0.82) for Xpert Bladder Cancer, and 0.75 (0.61-0.86) and 0.90 (0.84-0.94) for Bladder Epicheck. For ADXbladder and Uromonitor, meta-analyses incorporated four and six studies, respectively, resulting in pooled sensitivity and specificity values of 0.55 (0.40-0.69) and 0.60 (0.44-0.75) for ADXbladder, and 0.77 (0.61-0.88) and 0.96 (0.91-0.98) for Uromonitor.

**Conclusions:** This meta-analysis reveals that commercially UBTs for BC recurrence have varying diagnostic accuracy. Among the evaluated tests, Uromonitor demonstrated the highest pooled sensitivity and specificity, while Xpert Bladder Cancer and Bladder Epicheck showed reliable diagnostic performance. Further research is needed particularly for less extensively studied assays to establish their diagnostic performance.

## Introduction

Non-muscle-invasive bladder cancer (NMIBC) is characterized by a high recurrence rate ranging from 60% to 80% especially in patients with high-risk disease [1]. As a result, long-term surveillance is necessary, most commonly via cystoscopy, which is invasive, expensive and associated with moderate to severe discomfort in over half of patients. [2], [3], [4]. While urine cytology can be useful, it lacks sensitivity and is not routinely recommended in clinical guidelines [2]. To address these challenges, a number of urinary biomarker tests (UBTs) have been developed in recent years, including Bladder Epicheck (Nucleix; Rehovot, Israel), ADXbladder (Arquer Diagnostics; Sunderland, UK), Uromonitor (U-Monitor; Porto, Portugal), Xpert bladder cancer (Cepheid; Sunnyvale, California, USA) and Cxbladder Monitor. These tests aim to improve detection of recurrence while reducing the need for repeated cystoscopies [5]. However, none are currently approved by the FDA or recommended for routine clinical use [6].

While Xpert BC and Uromonitor have been individually studied, comprehensive and comparative meta-analyses comparing multiple commercially available UBTs using standardized methods remain limited. This gap limits clinicians’ ability to make informed choices between tests based on direct evidence. Additionally, certain promising tests like Cxbladder Monitor remain understudied with too few eligible studies available to conduct a meta-analysis, thereby restricting insights into their potential diagnostic utility. There is a need to deal with the high degree of heterogeneity across studies, including differences in inclusion criteria, tumour grade and risk stratification, previous use of intravesical therapies such as BCG, varying definitions of recurrence, inconsistent cut-off thresholds and differences in study designs. This variability complicates efforts to generalize findings or produce reliable pooled estimates. Moreover, most existing studies emphasize high-grade tumors, resulting in limited evidence on test performance in detecting low-grade recurrences, which are also clinically important in the surveillance of BC.

In the present study, we assessed the diagnostic performance of five commercially available UBTs (Xpert Bladder Cancer, Bladder Epicheck, ADXbladder, Uromonitor and CxBladder Monitor) in monitoring BC recurrence using a framework of meta-analysis of diagnostic test accuracy.

## Materials and methods

We followed the Preferred Reporting Items for Systematic Reviews and Meta-analyses of diagnostic test accuracy studies (PRISMA-DTA) guidelines (Supplementary Table 1) [7]. The study protocol was prospectively registered in the International Prospective Register of Systematic Reviews (PROSPERO) - (ID: CRD42024532700).

### Identification of novel commercially available urine-based biomarker tests

In the context of our work with the Cancer Incidence and Surveillance Modeling Network (CISNET) Bladder Cancer Modeling site, we developed and maintained an Evidence Map of the bladder cancer literature. Using this Evidence Map we identified tests that involve genomic, metabolomic or other-omic signatures, analyses on cells or cellular debris in urine. With input from clinician stakeholders and an advisory board, we prioritized for assessment 5 tests that are currently commercially available in the US. These index tests were Xpert® Bladder Cancer Monitor [8], ADXbladder™ [9], Bladder EpiCheck® [10], Uromonitor® [11], and Cxbladder Monitor® [12].

### Eligibility criteria

Eligible were studies including bladder cancer patients with NMIBC undergoing surveillance for recurrence, in which at least one of the five index tests was assessed against a reference standard with respect to diagnosing bladder cancer recurrence. We considered all studies in which we could extract the cross-classification of index and reference tests’ results about recurrence. We placed no restrictions on study design. We included both diagnostic cohorts and case-control designs, and for studies comparing more than one index tests, both parallel and paired designs. Eligible studies used cystoscopy-based surveillance, without restrictions on cystoscopy type (e.g., blue vs white light) or cadence.

### Data sources

We searched PubMed and Embase until May, 2025, without language and time restrictions. We used the commercial name of the tests (e.g. ADXbladder) along with the broad term “bladder” to identify relevant studies. Screening of the records was performed independently by two reviewers (K.T. and EEN) using the Rayyan software [13], first by screening titles and abstracts, followed by full-text screening.

During piloting, it was noted that multiple publications reporting on the same study populations were common. To identify and eliminate multiple publications of the same study population, we screened for overlapping authorship, institutions, study populations and sample sizes. When multiple records appeared to originate from the same study cohort, we examined them closely by cross-checking participant characteristics, study design, recruitment periods, outcomes reported and reference citations. In case of overlap, we retained the most complete and recent publication or the version with the most relevant data for our analysis.

### Data extraction

Two reviewers (K.H.T. and D.B.) independently extracted detailed data from included studies: study information (e.g. first author, publication year and study design), patient characteristics (e.g. country, sample size, number of recurrence cases, T stage, grade type), index test (e.g. follow-up time, threshold of test) and study results (e.g. sensitivity, specificity, positive predictive value (PPV), negative predictive value (NPV), true positive (TP), true negative (TN), false positive (FP), and false negative (FN)). When there are multiple positivity cutoffs/thresholds, we extracted 2 by 2 tables for each cutoff. Finally, each diagnostic test included the following measures: sensitivity (95% CI), specificity (95% CI), and TP, TN, FP, and FN for each threshold.

We categorized events as either recurrence only or composite events (recurrence and progression) based on each study’s definitions.

### Risk of bias assessment

The risk of bias was assessed according to the updated Quality Assessment of Diagnostic Accuracy Studies tool (QUADAS-2) by two reviewers (K.H.T and EEN) [14]. This allowed for a critical assessment of the quality of the included studies and their potential bias. Each study was therefore judged by a set of questions addressing the following areas of bias: 1) Patient Selection, 2) Index Trial, 3) Reporting Standard, and 4) Flow and Timing. Arriving at a conclusion per category that judged the risk of bias as “Low”/ “High”/ “Unclear”. The plots regarding the QUADAS-2 results were extracted by RevMan [15].

## Statistical analyses

The bivariate Generalized Linear Mixed Model (GLMM) was applied to synthesize diagnostic accuracy data [16], [17]. Additionally, the random effects model was also fitted using the glmer function with a binomial logit link [18]. The meta-analyses were based on a dataset containing diagnostic test performance data, including 95% CI sensitivity, 95% CI specificity, TP, TN, FP, and FN. The diagnostic meta-analysis models estimated the following 5 parameters: mean sensitivity, mean specificity, the SDs of the random effects of sensitivity and specificity, and the correlation between the random effects of sensitivity and specificity. A subgroup analysis was performed based on tumor grade. Results were then presented using forest plots for sensitivity and specificity with 95% confidence intervals and the summary receiver operating characteristic (sROC) curve for each diagnostic test [19]. Meta-analyses and summary ROC curves were conducted in R language (version 4.4.2) on R studio [20] using the “lme4” and “pROC” packages respectively.

## Results

A total of 307 studies were initially identified through the literature search across databases. The number of records for each UBT are presented in details in the PRISMA flow charts (Supplementary Figures 1-6). Following the title and abstract screening, 143 records remained to be assessed for eligibility based on the full-text article. Studies that were excluded after this step are presented in Supplementary Table 2 and flow charts (Supplementary Figures 1-6). Finally, 36 studies met the inclusion criteria, assessing the diagnostic accuracy of UBTs for detecting BC recurrence.

### Study characteristics

Table 1 provides characteristics of the included studies. The number of recurrence cases ranged from 40 to 1,431 patients, with a total of 10,556 participants across all included studies. The median duration of follow-up ranged from 2 to 48 months. Cystoscopy and histological examination were used as reference methods in the studies. Most studies defined recurrence as cystoscopy-and histologically-confirmed recurrence of NMIBC without reference to disease progression. A smaller group of studies included composite recurrence endpoints, where recurrence was represented by Ta, T1, CIS stages, while progression was typically defined as T2 stage and G3 grade. The majority of studies were prospective designs without consecutive sampling.

**Table 1.**
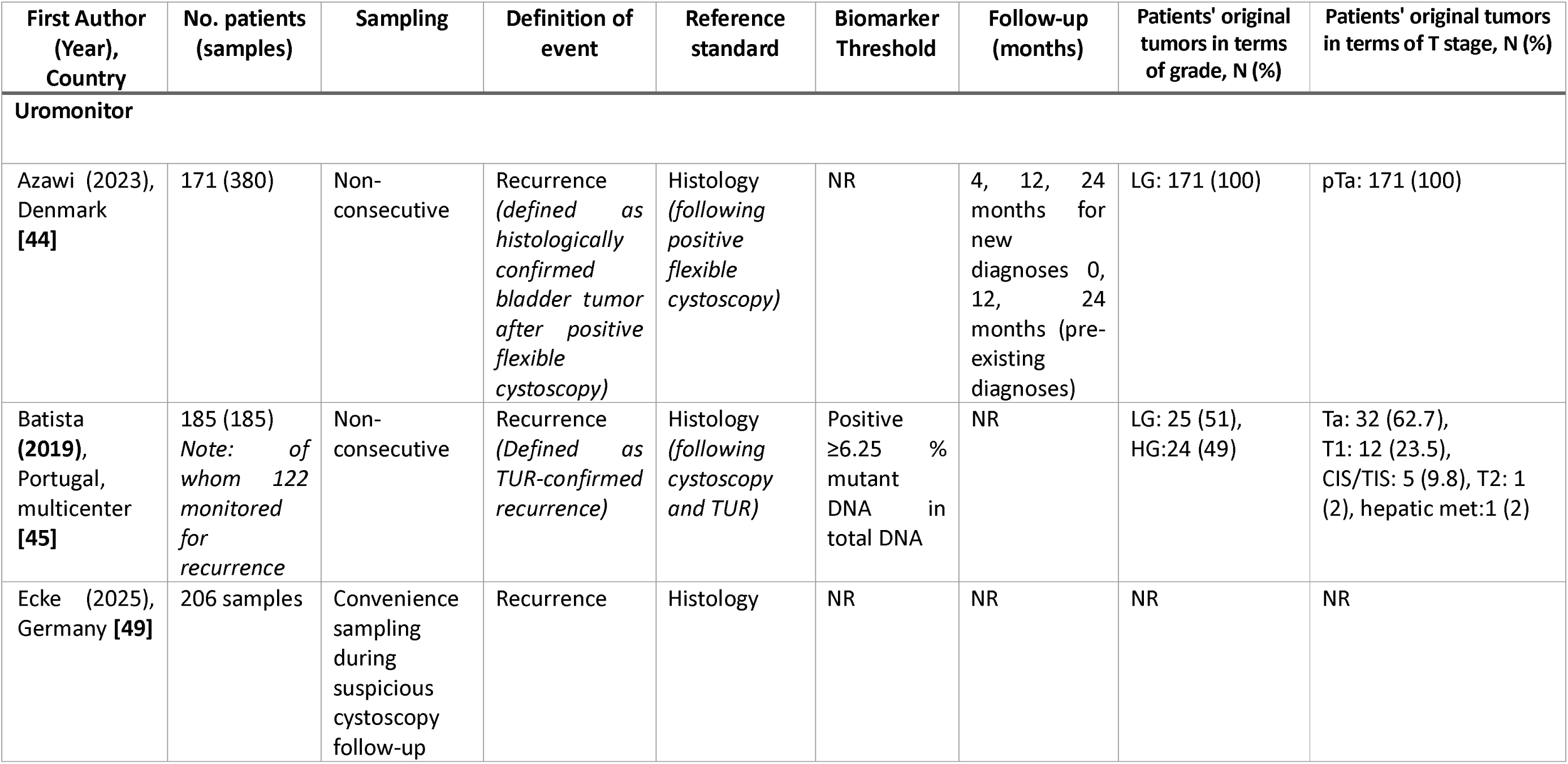
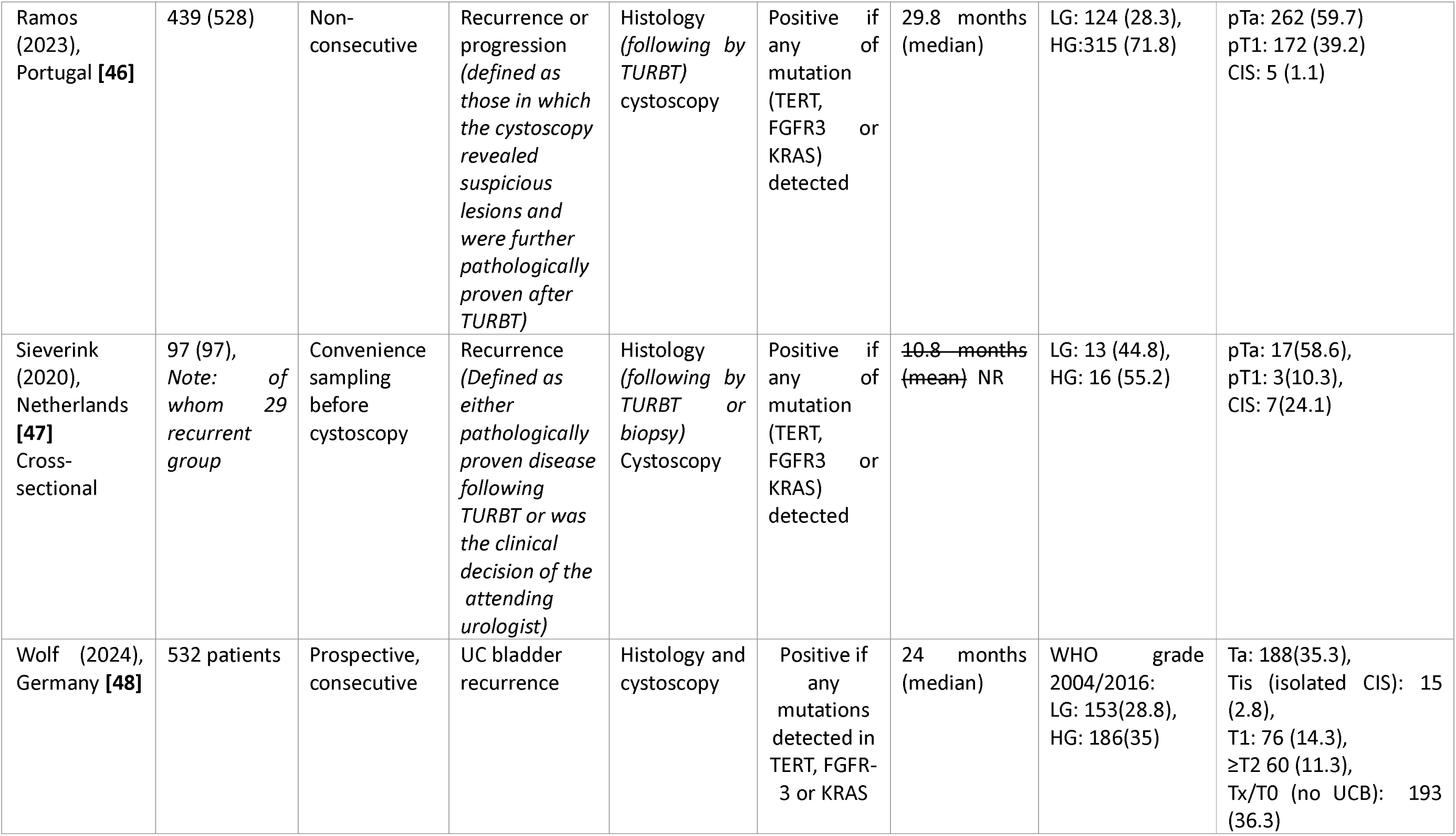
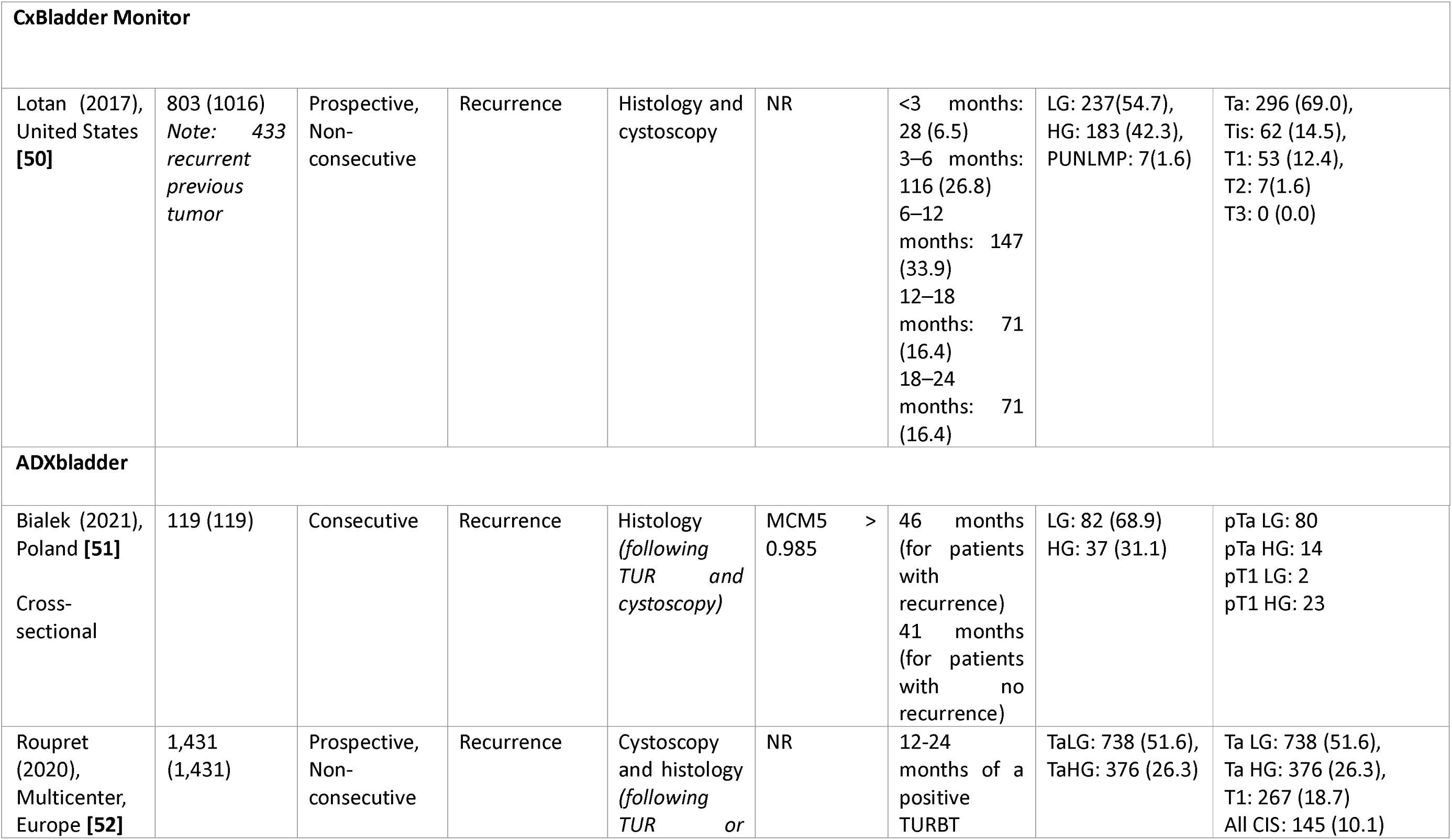
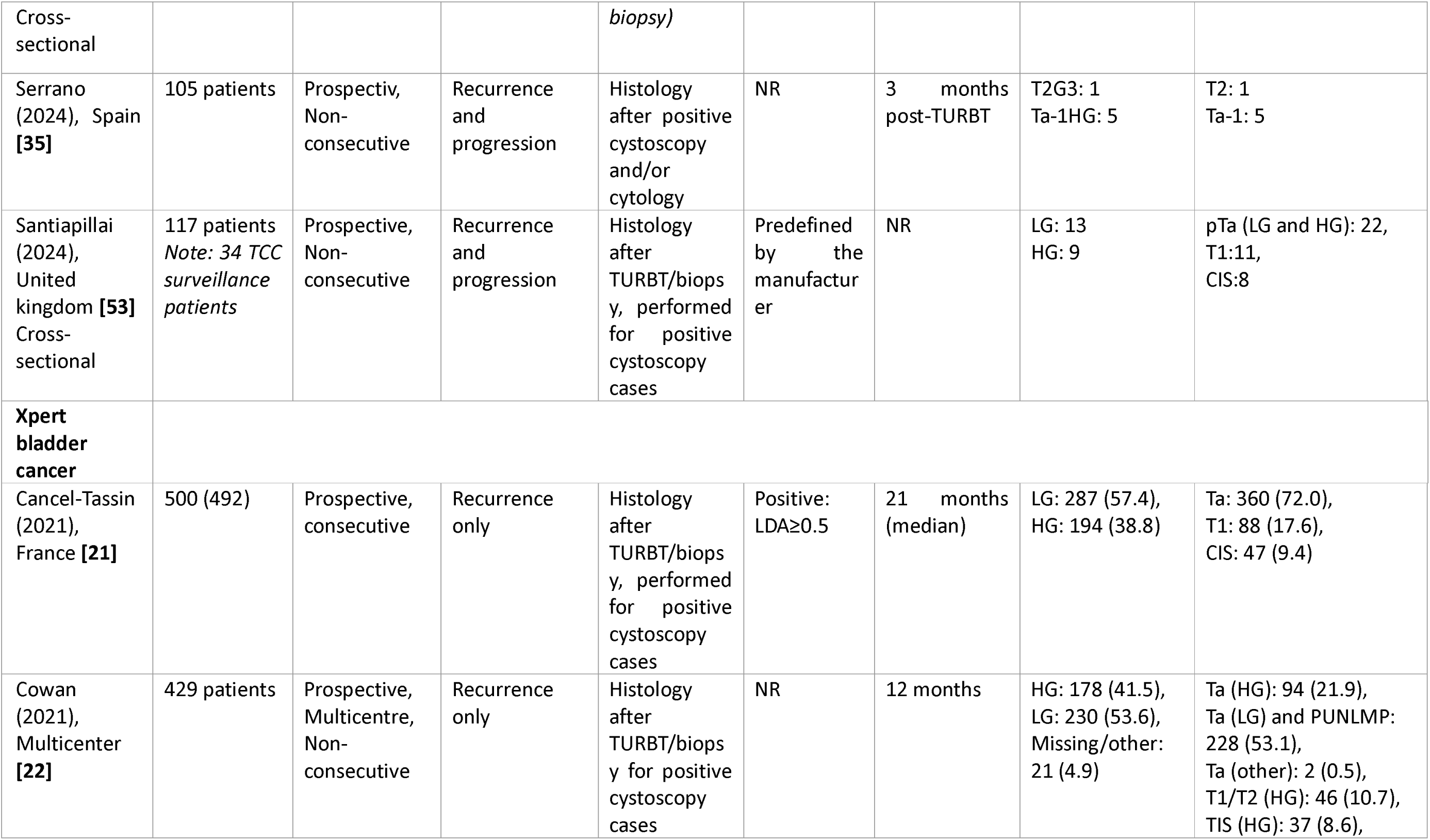
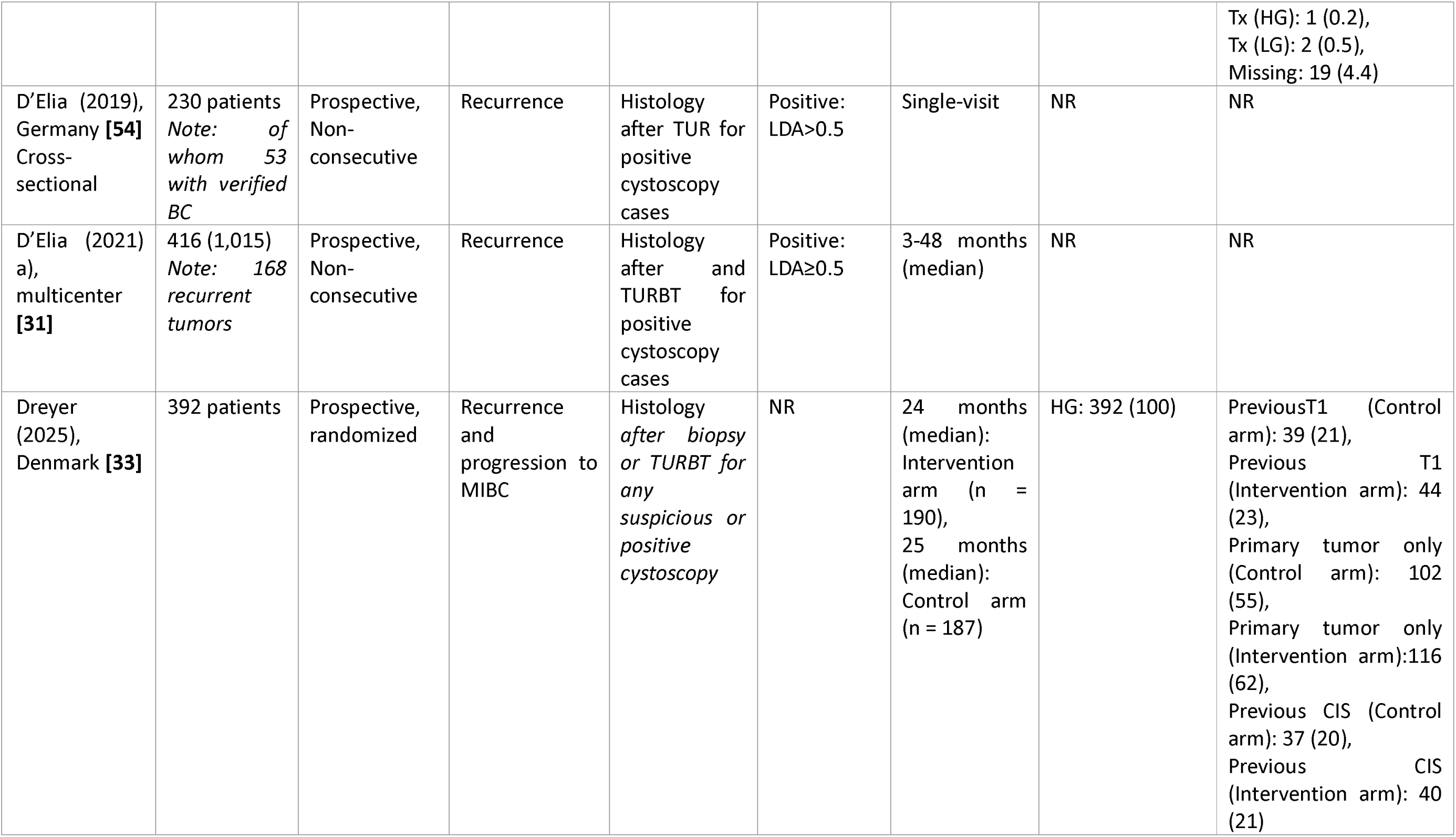
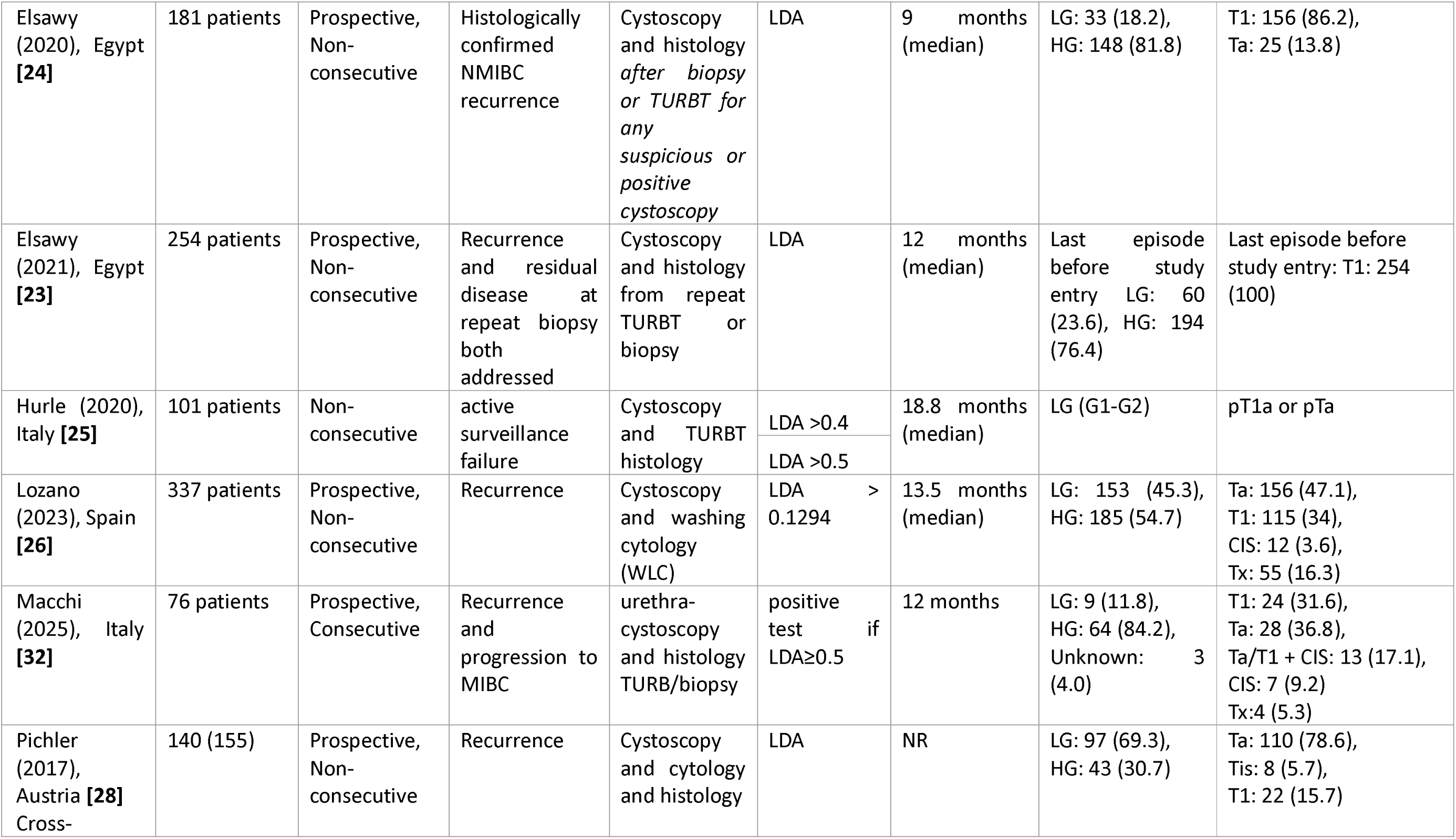
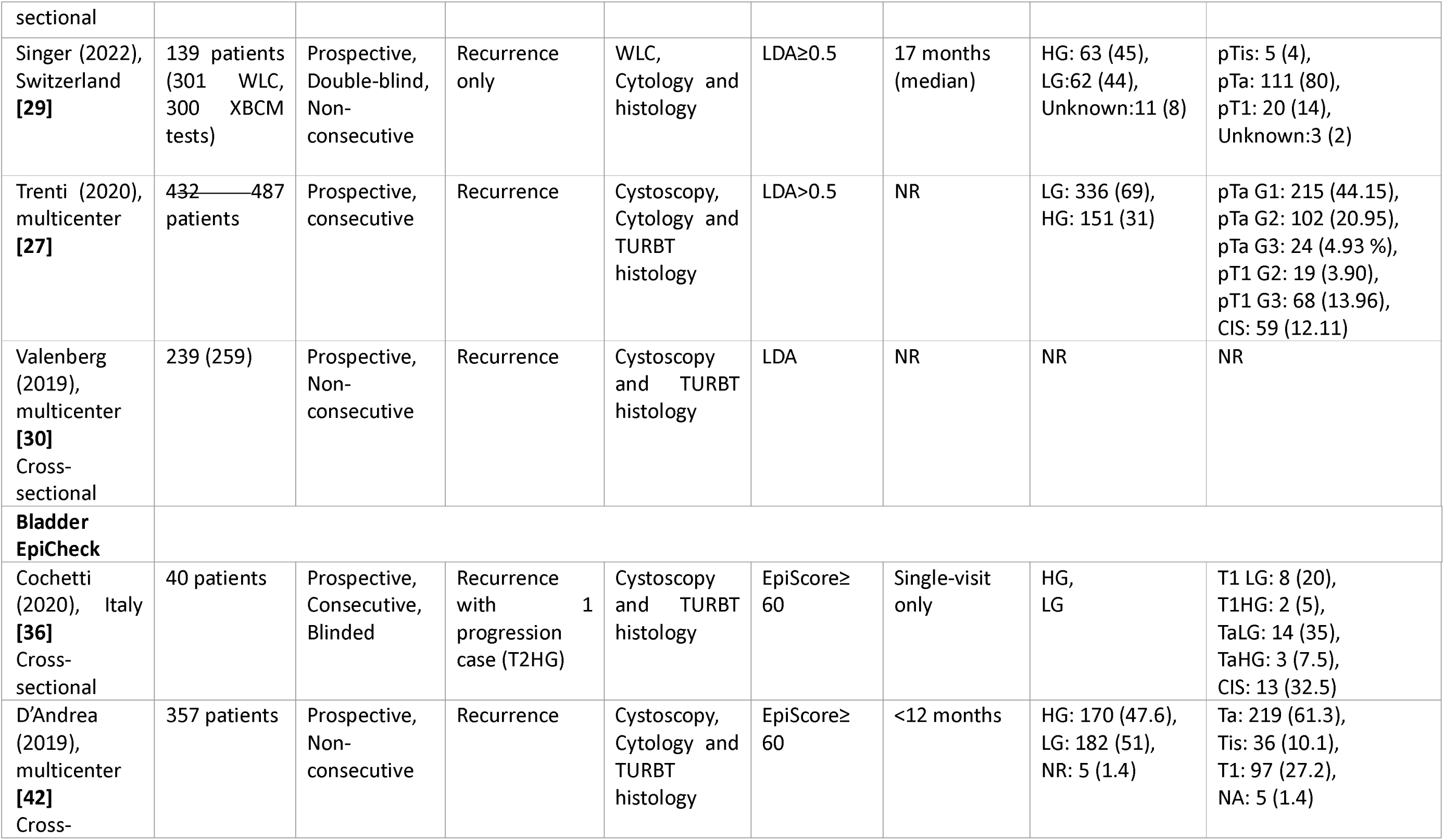
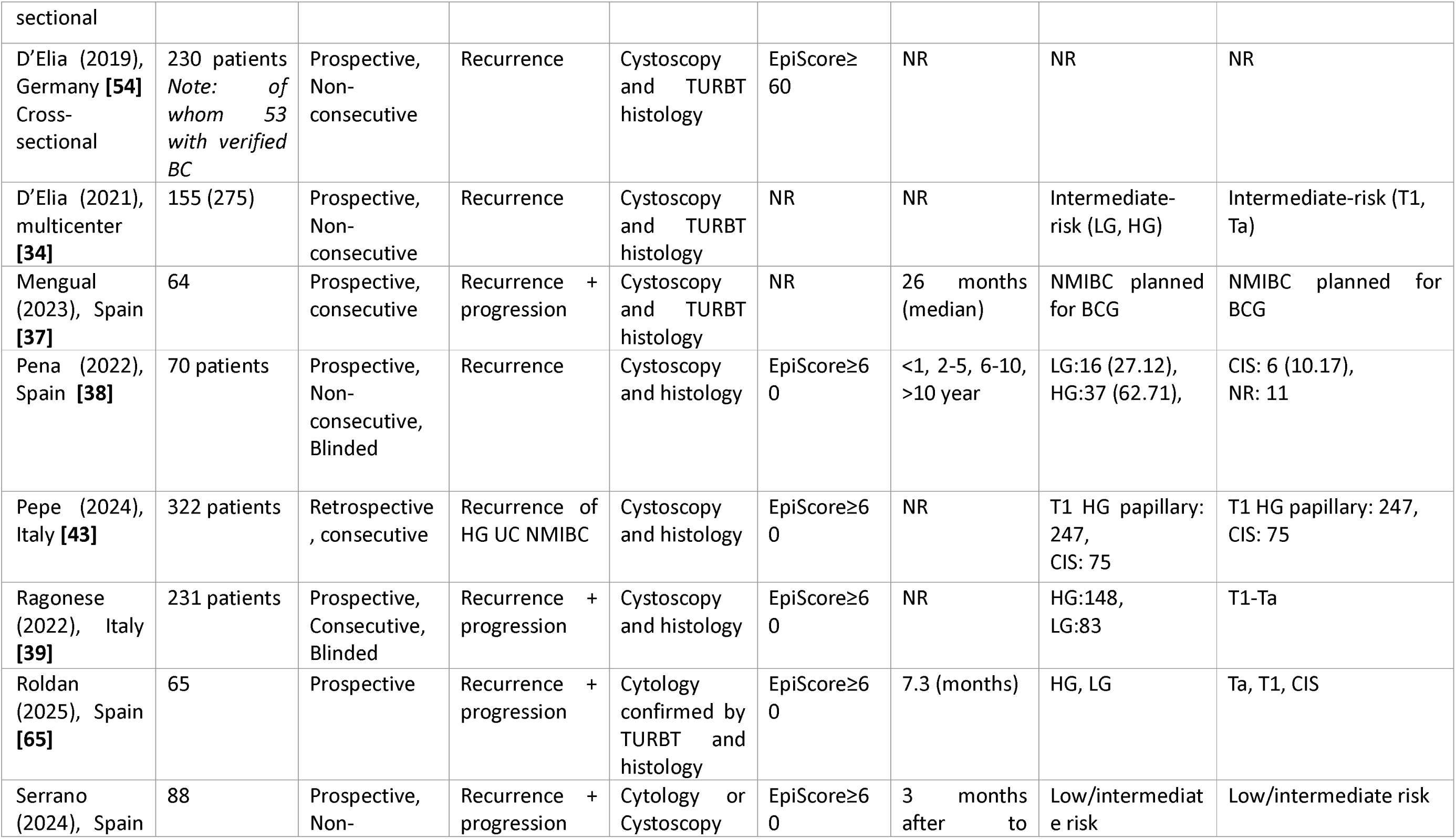
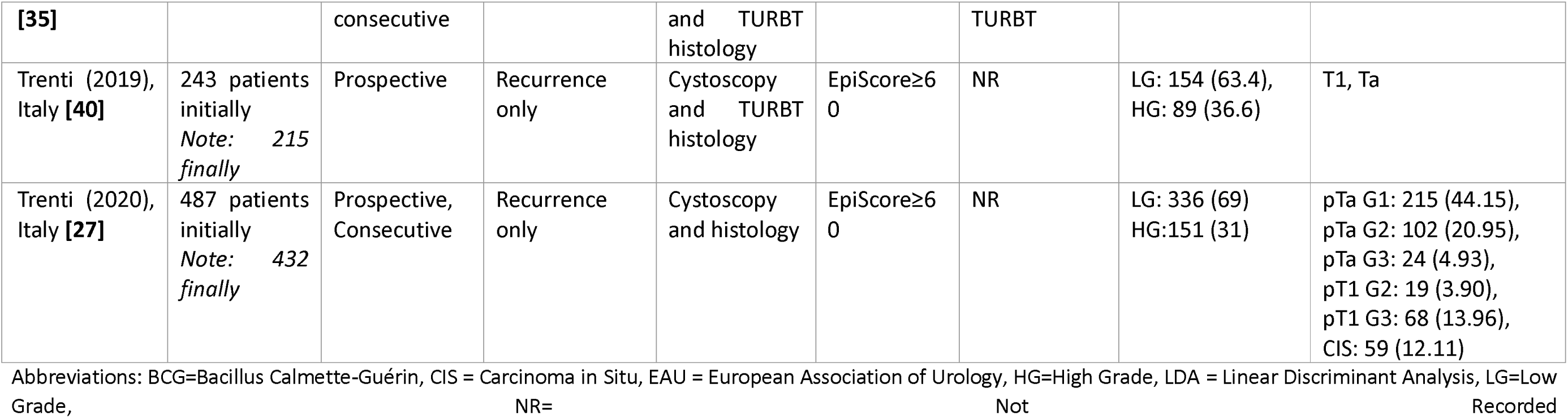
Characteristics of included studies in the analysis for diagnostic estimates of BC recurrence.

The reported sensitivities and specificities ranged from 50-100% and 33-99%, respectively. Thirteen studies used the Xpert Bladder Cancer Monitor as a diagnostic tool for recurrence [21], [22], [23], [24], [25], [26], [27], [28], [29], [30], [31], [32], [33]. Ten studies analysed Bladder Epicheck for monitoring patients with bladder cancer [34], [35], [36], [37], [38], [39], [40], [41], [42], [43]. Six studies provided data on the diagnostic values of Uromonitor for diagnosing recurrence during bladder cancer surveillance [44], [45], [46], [47], [48], [49]. One study analysed the CxBladder Monitor for diagnosing recurrence during bladder cancer follow-up [50]. Four studies evaluated the diagnostic accuracy of ADXbladder for detecting recurrence [35], [51], [52], [53]. The included studies from each test that were used in meta-analyses are presented in Table 2, providing the overall diagnostic estimates of the test for the entire population.

**Table 2.** The individual studies included in the meta-analysis of overall grade, including the results concerning with HG and/or LG, for each diagnostic test.

| First author<br>(year) | Sensitivity %<br>(95 % CI) | Specificity %<br>(95 % CI) | TP | TN | FP | FN |
| --- | --- | --- | --- | --- | --- | --- |
| **Uromonitor** |  |  |  |  |  |  |
| Azawi (2023)<br>[44] | 89.7 (75.8–<br>97.1) | 96.2 (93.6–98) | 35 | 328 | 13 | 4 |
| Batista (2019)<br>[45] | 73.5 | 93.2 | 25 | 82 | 6 | 9 |
| Ecke (2025)<br>[49] | 57.4 | 94.9 | 63 | 73 | 4 | 47 |
| Ramos (2023)<br>[46] | 87.2 | 99.2 | 41 | 478 | 3 | 6 |
| Sieverink<br>(2020) [47] | 93.1 (75.8–<br>98.8) | 85.4 (75.8–<br>93.4) | 27 | 41 | 7 | 2 |
| Wolff (2024)<br>[48] | 49.1 | 93.3 | 141 | 154 | 11 | 146 |
| **ADXbladder** |  |  |  |  |  |  |
| Bialek (2021)<br>[51] | 73.5 (62.7–<br>82.6) | 33.3 (18.6- 51) | 61 | 12 | 24 | 22 |
| Roupret<br>(2020) [52] | 44.9 (36.1-54) | 71.1 (68.5-73.5) | 57 | 927 | 377 | 70 |
| Santiapillai<br>(2024) [53] | 33.3 (0.80-90.5) | 54.8 | 1 | 17 | 14 | 2 |
| Serrano<br>(2024) [35] | 45.4 | 74.0 | 5 | 57 | 20 | 6 |
| **Xpert Bladder**<br>**Cancer** |  |  |  |  |  |  |
| Cancel-Tassin<br>(2021) [21] | 72.7 (58.0 -<br>83.7) | 73.7 (69.4 -<br>77.5) | 32 | 330 | 118 | 12 |
| Cowan (2021)<br>[22] | 60.3 (47.5–<br>71.9) | 76.5 (72.0–<br>80.6) | 35 | 284 | 87 | 23 |
| D’Elia (2021)<br>[31] | 52.4 | 78.4 | 88 | 664 | 183 | 80 |
| Dreyer (2025)<br>[33] | 91 (71 – 99) | 65 (59 – 71) | 20 | 109 | 59 | 2 |
| Elsawy (2020)<br>[24] | 73.7 (67-79) | 79.6 (74-84) | 14 | 129 | 33 | 5 |
| Elsawy (2021)<br>[23] | 85.9 (82-89) | 72.3 (68-76) | 85 | 112 | 43 | 14 |
| Hurle (2020)<br>[25] | 30 | 90.2 | 18 | 37 | 4 | 42 |
| Lozano (2023)<br>[26] | 69.4 | 68.8 | 34 | 198 | 90 | 15 |
| Macchi (2025)<br>[32] | 88.9 | 66.7 | 16 | 22 | 11 | 2 |
| Pitchler (2017) [28] | 84.0 (69 - 93) | 91.0 (83 - 96) | 36 | 88 | 9 | 7 |
| Singer (2022) [29] | 58.0 (50-65) | 89.0 (83-94) | 29 | 224 | 27 | 21 |
| Trenti (2020) [27] | 66.3 (55.7-75.8) | 76.5 (71.6-80.9) | 61 | 260 | 80 | 31 |
| Van Valenberg (2019) [30] | 74.0 (60.0-85.0) | 80 (73.0-85.0) | 32 | 156 | 40 | 11 |
| **Bladder EpiCheck** |  |  |  |  |  |  |
| Cochetti (2020) [36] | 100 (82.4-100) | 90.9 (72.2-98.4) | 18 | 20 | 2 | 0 |
| D'Andrea (2019) [42] | 67.3 (52.0-80.0) | 88.0 (84.0-91.0) | 33 | 271 | 37 | 16 |
| D'Elia (2021) [34] | 48.9 | 83.5 | 22 | 192 | 38 | 23 |
| Mengual (2023) [37] | 85 | 78 | 11 | 40 | 11 | 2 |
| Pepe (2024, LG) [43] | 100 (63.1 - 100) | 99 (97 - 99.8) | 8 | 331 | 3 | 0 |
| Pepe (2024, HG) [43] | 83.3 (58.6 - 96.4) | 98.6 (96.5 - 99.6) | 15 | 300 | 4 | 3 |
| Pena (2022) [38] | 91.7 | 91.9 | 22 | 44 | 4 | 2 |
| Ragonese (2022) [39] | 80.0 | 84.0 | 57 | 116 | 22 | 14 |
| Serrano (2024) [35] | 27.2 | 90.9 | 3 | 70 | 7 | 8 |
| Trenti (2019) [40] | 62.3 (32.6-59.7) | 86.3 (79.6-91.4) | 43 | 126 | 20 | 26 |
| Trenti (2020) [27] | 64.2 (53.5-73.9) | 82.1 (77.6-85.6) | 59 | 279 | 61 | 33 |

Abbreviations: CI=Confidence Interval, FN= False Negative, FP=False Positive, TN=True Negative, TP=True Positive

Four studies included in this systematic review evaluated more than one index test: Serrano et al. [35] provided results for ADXbladder and Bladder Epicheck, reporting diagnostic performance metrics for both tests in parallel. Moreover, three studies assessed both Xpert Bladder Cancer Monitor and Bladder Epicheck, allowing a head-to-head comparison [34], [41], [54].

Across the included studies, follow-up strategies for assessing recurrence varied. The majority of studies (n=20) implemented longitudinal follow-up. Fiftheen studies do not report any follow-up duration and four studies presented only one follw-up time-point from a single visit.

### Quality assessment

#### Xpert Bladder Cancer

The QUADAS-2 results for Xpert Bladder Cancer are presented in Supplementary Figures 7-8. Ten studies presented unclear risk of bias regarding patient selection, because either the sample was not consecutive or random or because they did not avoid inappropriate exclusions [21], [22], [23], [24], [25], [26], [28], [29], [30], [31]. Regarding the index test, the risk of bias was deemed as unclear for 7 studies out of 13, due to studies that did not mention blinding of lab staff performing Xpert Bladder Cancer, cystoscopy or TURBT results [23], [24], [25], [27], [28], [30], [31].

Seven studies also were judged as of unclear risk of bias as regards the reference standard due to the absent statement of whether clinicians such as urologists or pathologists were blinded [24], [25], [28], [32], [33], [34], [41]. Seven studies were judged as of high-risk of bias in the flow and timing domain because not all patients were included in the analysis, not all of them received a reference standard or the same reference standard [22], [25], [26], [28], [29], [30], [41]. Three studies also were judged as of unclear risk of bias in the same domain ([21], [23], [24]) due to the fact that the analysis did not use all patients. Finally, applicability concerns were low in all studies across the assessed domains.

#### Bladder Epicheck

The QUADAS-2 assessment of Bladder Epicheck studies is presented in Supplementary Figures 9-10. In the patient selection domain three studies were assessed as of high risk of bias ([34], [36], [42]), because either the sample was not consecutive or random or because they did not avoid inappropriate exclusions. The remaining studies were appraised as of unclear risk of bias in the same domain since it was not clarified if inappropriate exclusions were avoided. Regarding the index test, five studies were assessed as of unclear risk of bias due to studies that did not mention either if blinding was used when interpreting Bladder Epicheck results or whether the used threshold was pre-specified ([27], [34], [35], [37], [40]). As far as the reference standard was concerned, 8 studies out of 10 were judged as of unclear risk of bias due to either no report of clinicians being blinded to Bladder Epicheck results or because it was not clear if the reference standard was likely to correctly classify the target condition ([34], [35], [37], [38], [40], [41], [42], [43]). Moreover, four studies presented as of high risk of bias as regards the flow and timing domain ([42], [34], [40], [41]), because they did not use all patients in their analysis and not all patients received a reference standard or the same reference standard. Three studies were also assessed as of unclear risk of bias ([36], [37], [39]), because not all patients were included in their analysis. Finally, applicability concerns were low in all studies across the assessed domains.

#### ADXBladder

The QUADAS-2 assessment of ADXBladder studies revealed variable risk of bias judgements, while applicability concerns were low in all studies across the assessed domains (Supplementary Figures 11-12). Specifically, one study [51] was rated as high-risk in the patient selection domain because the study did not avoid case-control design and inappropriate exclusions. This study also presented as of high risk of bias as regards flow and timing domain due to not all included patients having received a reference standard and/ or the same reference standard and not all of them were used in the analysis. The remaining studies were appraised as of unclear risk of bias on patient selection due to inappropriate exclusions. Three studies were of unclear risk of bias on the index test ([35], [51], [53]), because they did not mention if lab technicians running the test were blinded to cystoscopy or pathology results. Risk of bias related to the reference standard was unclear in two studies ([35], [51], [53]) that did not explicitly state whether clinicians interpreting cystoscopy/histology were blinded. Finally, one study [52] presented as of unclear risk of bias regarding the flow and timing domain because not all patients were included in the analysis.

#### Uromonitor

The QUADAS-2 assessment of Uromonitor studies revealed unclear risk of bias across them (Supplementary Figures 13-14). Specifically, four studies ([45], [46], [47], [55]) presented as of unclear risk of bias in patient selection due to non-consecutive or non-random sampling and one study [48] presented as of high risk of bias because the included population was not restricted to NMIBC patients. One study [55] was judged as of unclear risk of bias for index test domain because it did not mention whether Uromonitor was performed blinded to the cystoscopy or histology results. Unclear risk was also presented as regards the reference standard for three studies ([46], [55], [56]) that did not report blinding of the clinicians interpreting the reference standard. Moreover, flow and timing was presented as of unclear risk of bias for four studies ([44], [45], [46], [47]) because not all patients were included in the corresponding analysis of each study. Most studies presented low applicability concern across all domains.

### Meta-analysis results

#### Diagnostic performance of Xpert Bladder Cancer Monitor

The meta-analysis of studies evaluating Xpert Bladder Cancer included 13 studies, as presented in Table 2. The pooled estimates for sensitivity and specificity with confidence intervals of the studies were 0.71 (0.61, 0.79) and 0.78 (0.74, 0.82), respectively (Figures 1 & 2). For sensitivity, point estimates ranged from 0.30 to 0.91, while specificity estimates ranged from 0.65 to 0.91. Notably, only a few studies had confidence intervals that overlapped substantially with the pooled estimates, suggesting variability in the data (Figure 3).

**Figure 1.**
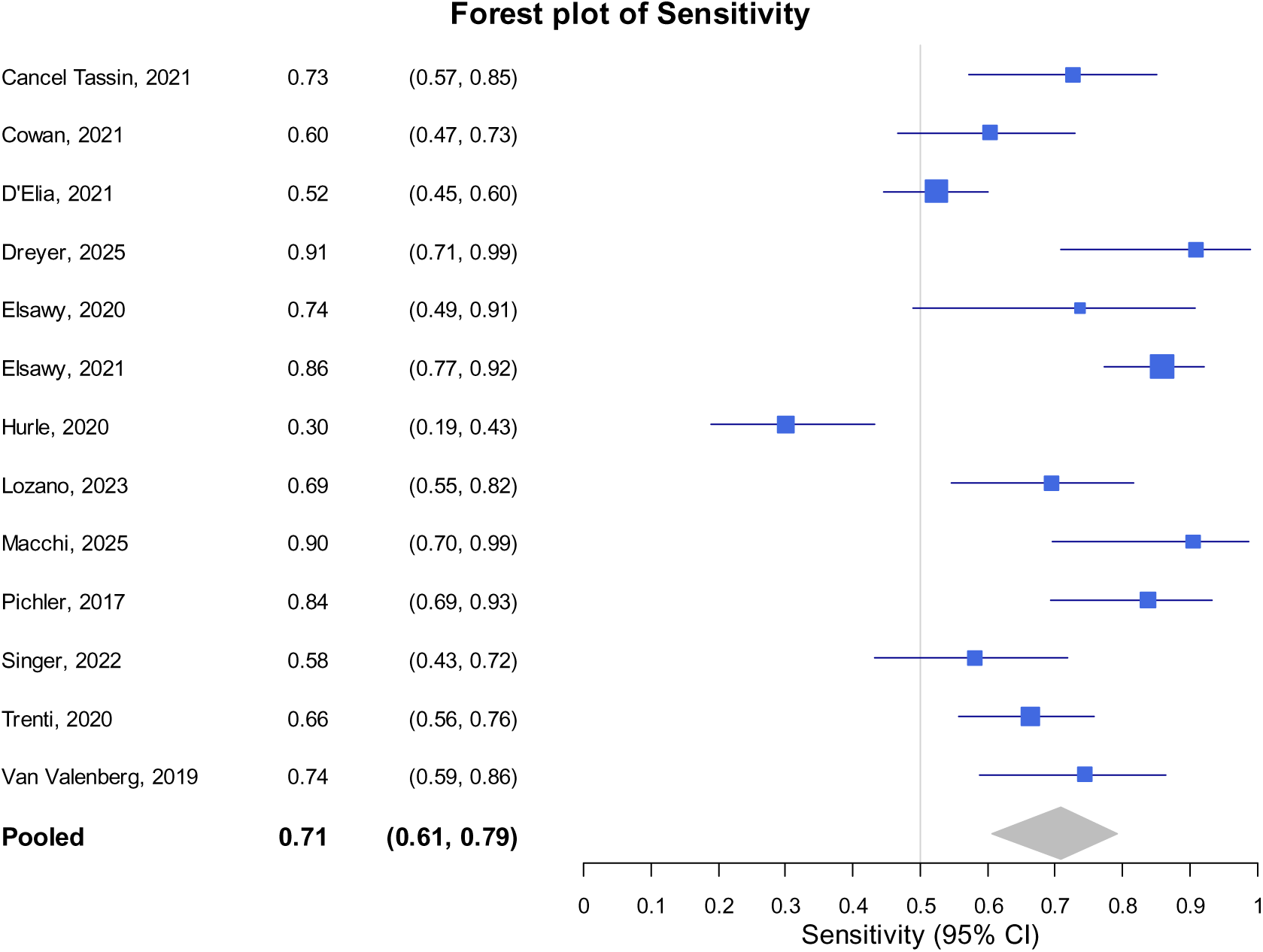
Forest plot of Xpert Bladder Cancer (sensitivity)

**Figure 2.**
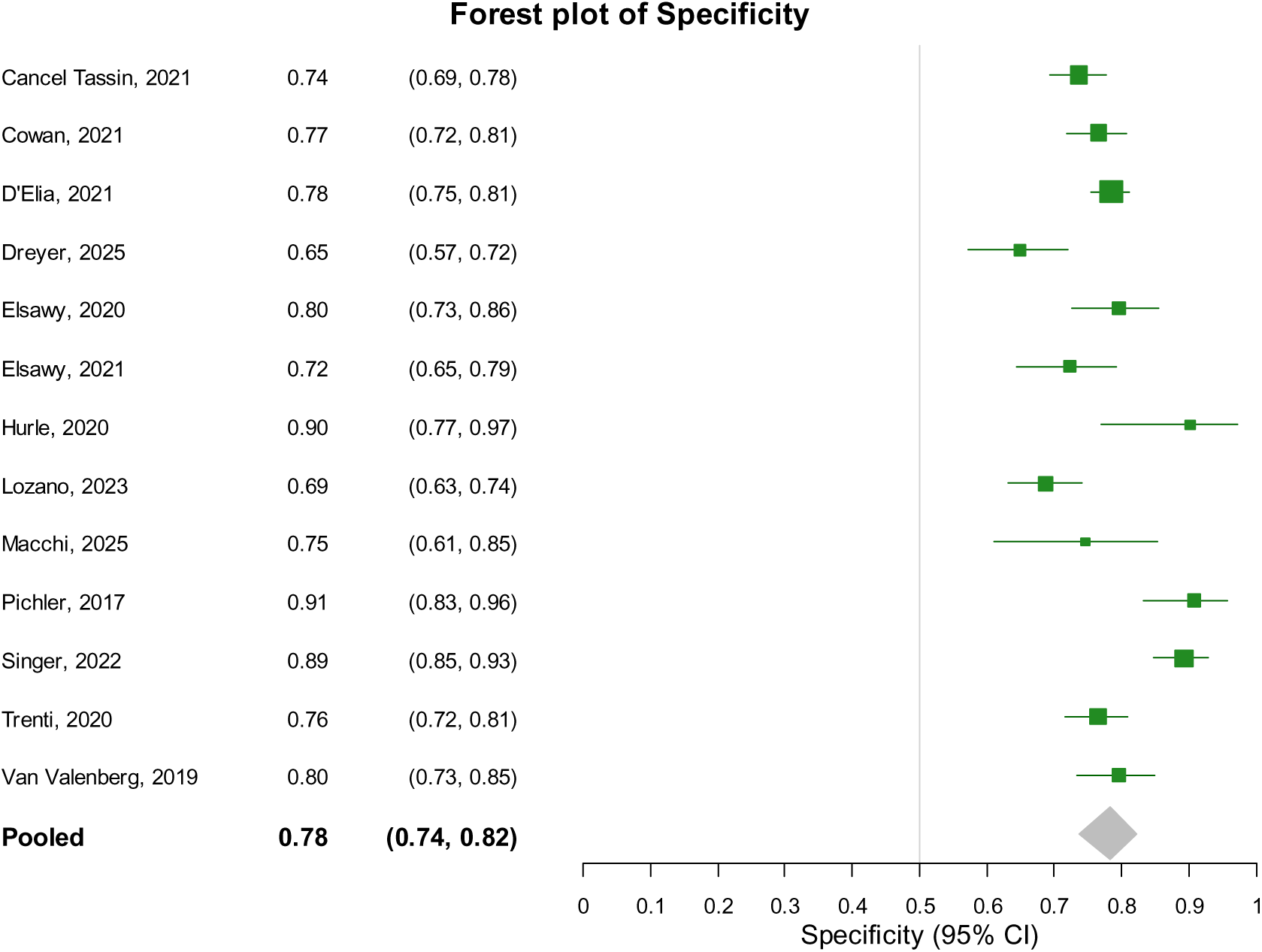
Forest plot of Xpert Bladder Cancer (specificity)

**Figure 3.**
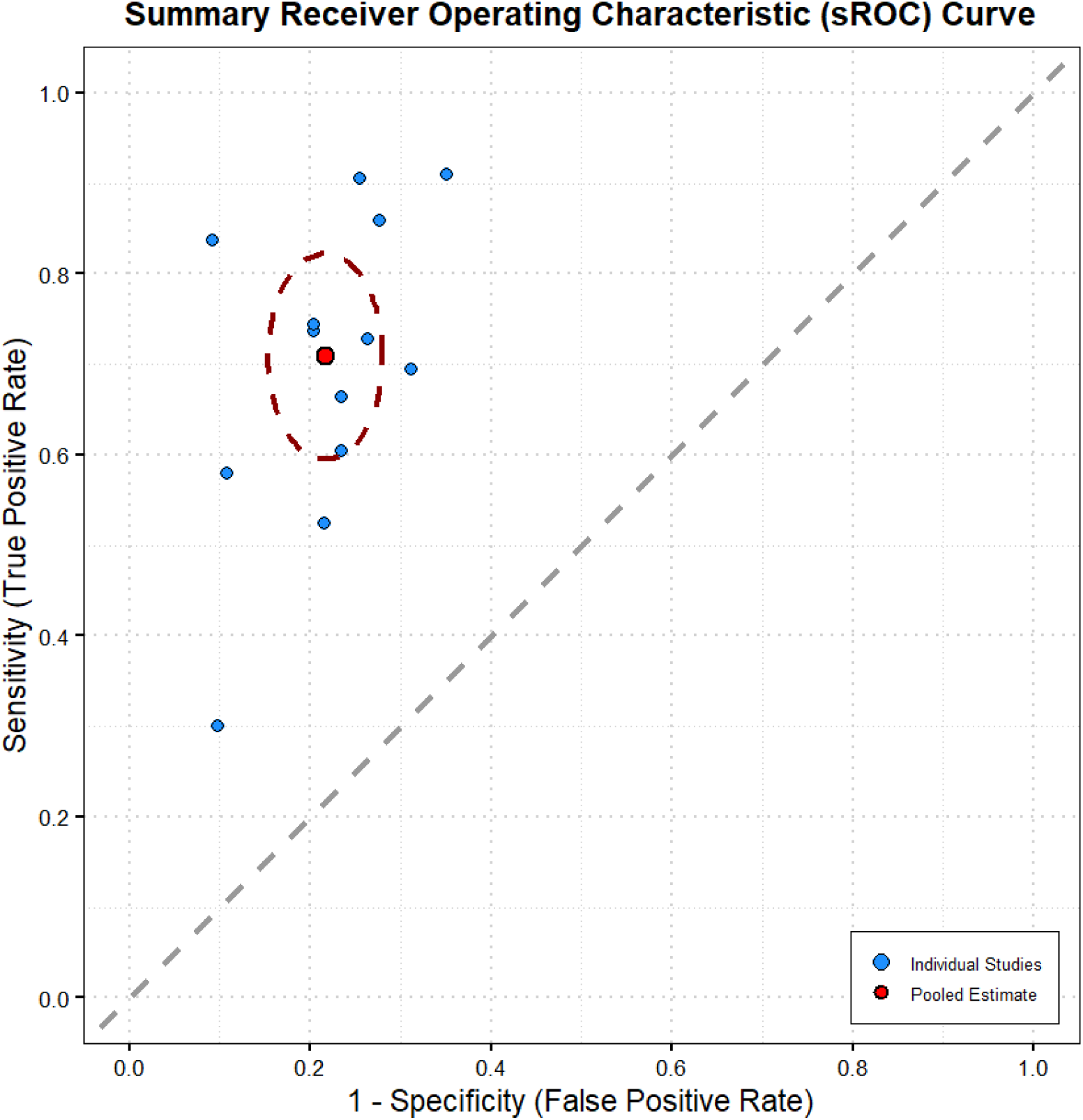
Summary sensitivity and specificity curve for Xpert Bladder Cancer

#### Diagnostic performance of Bladder Epicheck

The meta-analysis of studies regarding the Bladder Epicheck used 10 studies (Table 2). The pooled estimate of sensitivity and specificity with confidence intervals of the studies are 0.75 (0.61, 0.86) and 0.90 (0.84, 0.94), respectively (Figures 4 & 5). For sensitivity, point estimates ranged from 0.27 to 1.00, while specificity estimates ranged from 0.78 to 0.99 (Figure 6).

**Figure 4.**
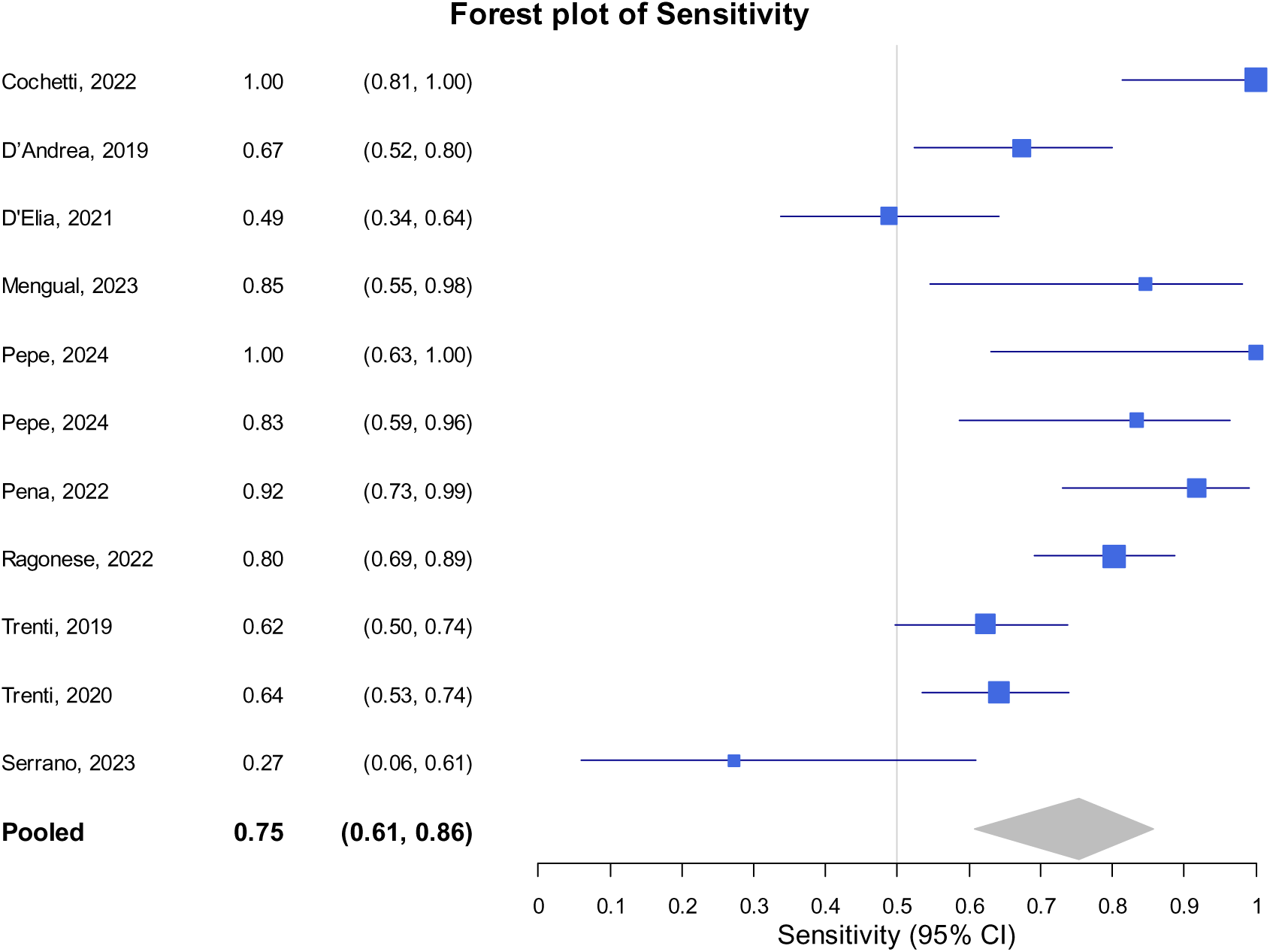
Forest plot of Bladder Epicheck (sensitivity)

**Figure 5.**
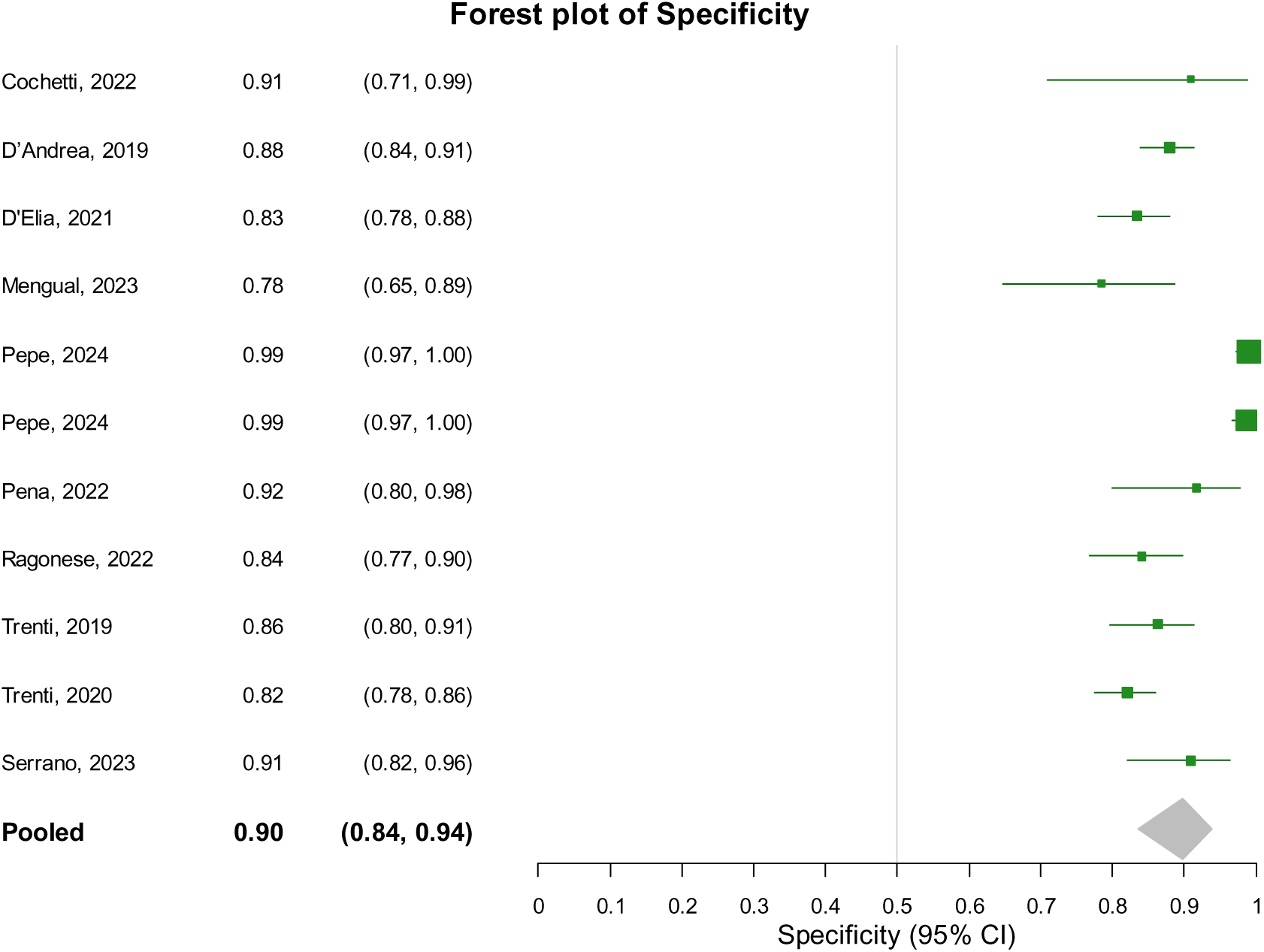
Forest plot of Bladder Epicheck (specificity)

**Figure 6.**
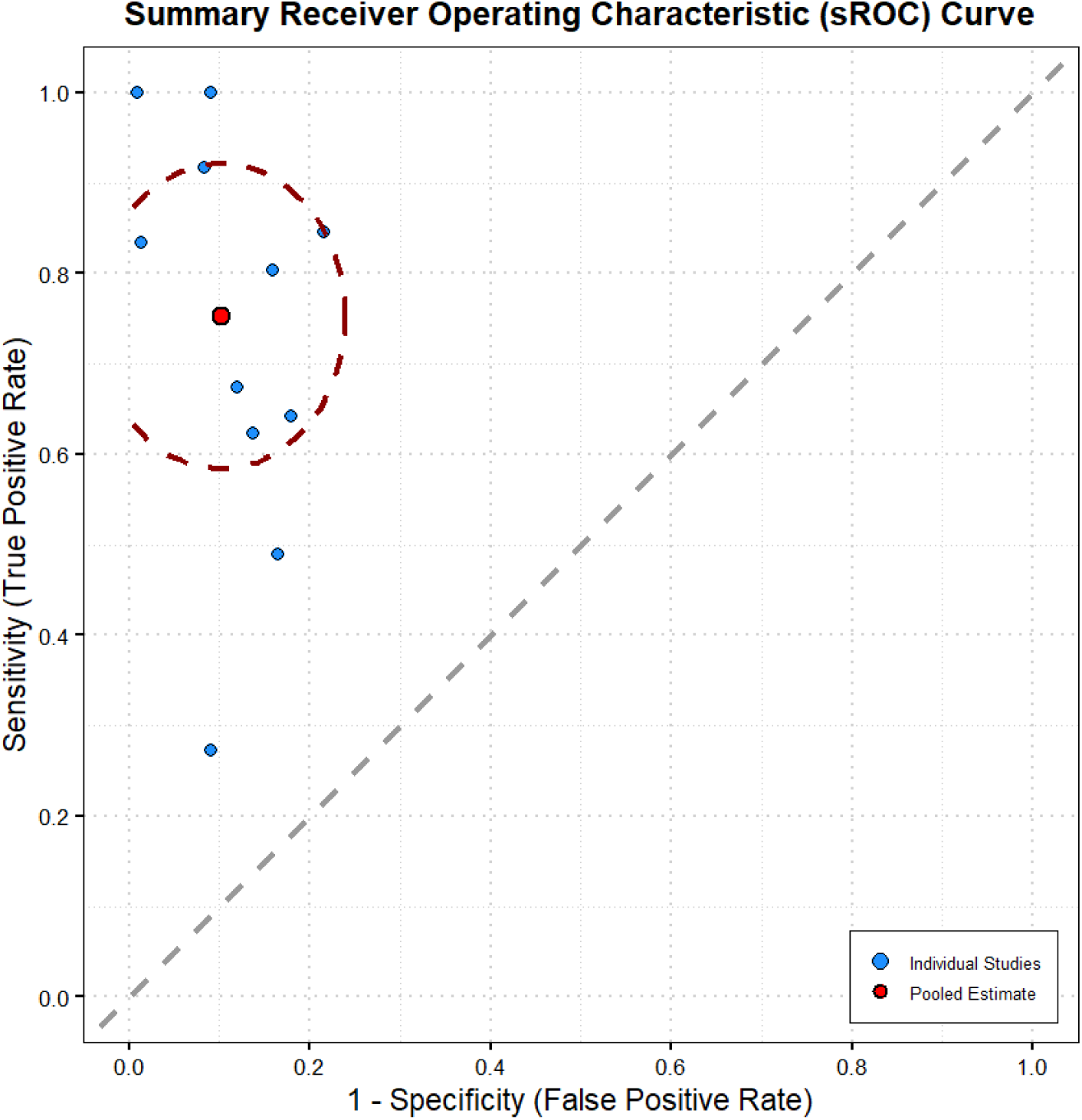
Summary sensitivity and specificity curve for Bladder Epicheck

#### Diagnostic performance of ADXBladder

Four studies were included in the meta-analysis regarding of ADXBladder (Table 2). The pooled estimate of sensitivity and specificity with confidence intervals of five studies was 0.55 (0.40, 0.69) and 0.60 (0.44, 0.75), respectively (Figures 7 & 8). Point estimates ranged from 0.45 to 0.73 for sensitivity and point estimates for specificity ranged from 0.33 to 0.74 (Figure 9).

**Figure 7.**
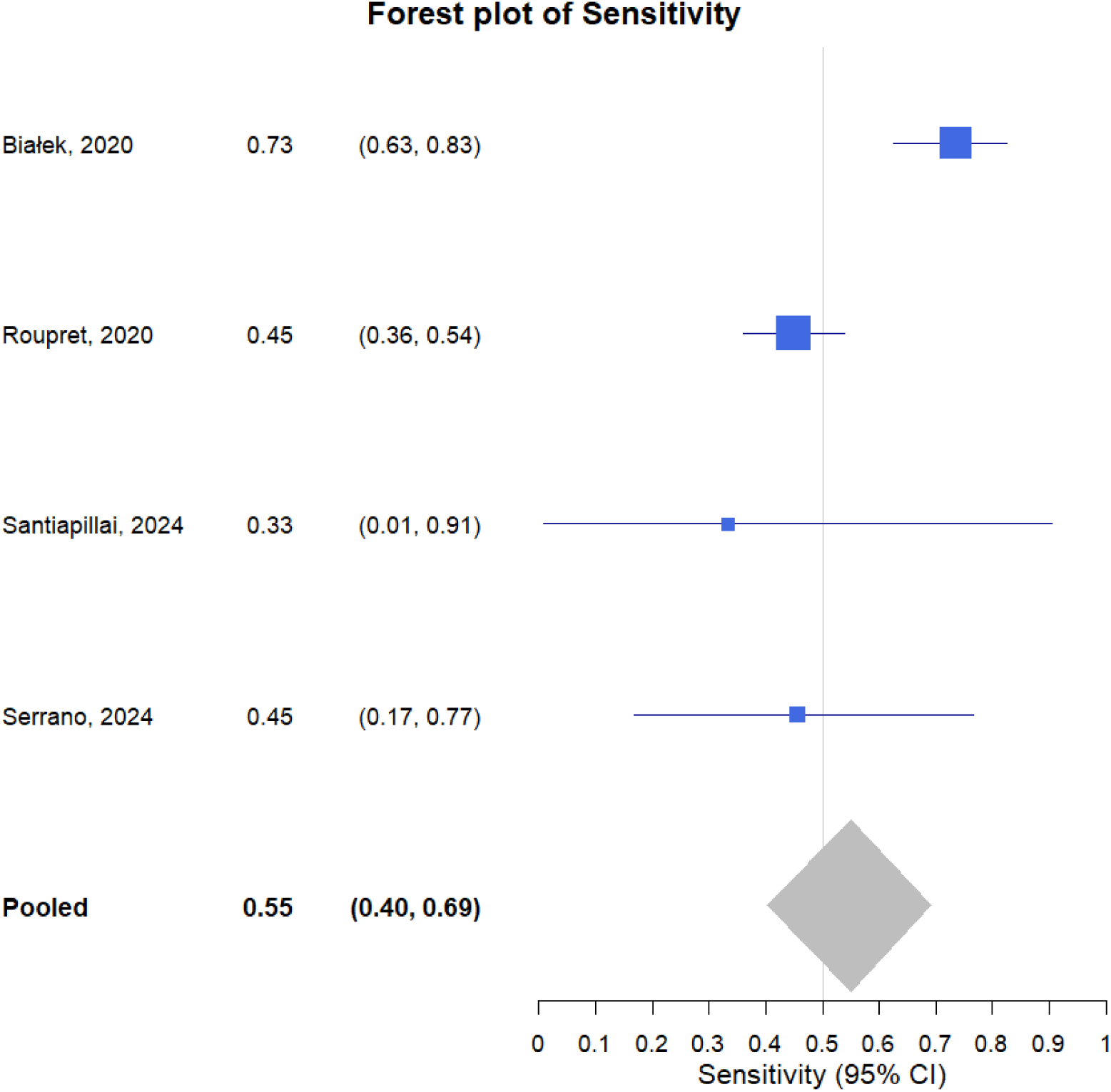
Forest plot of ADXbladder (sensitivity)

**Figure 8.**
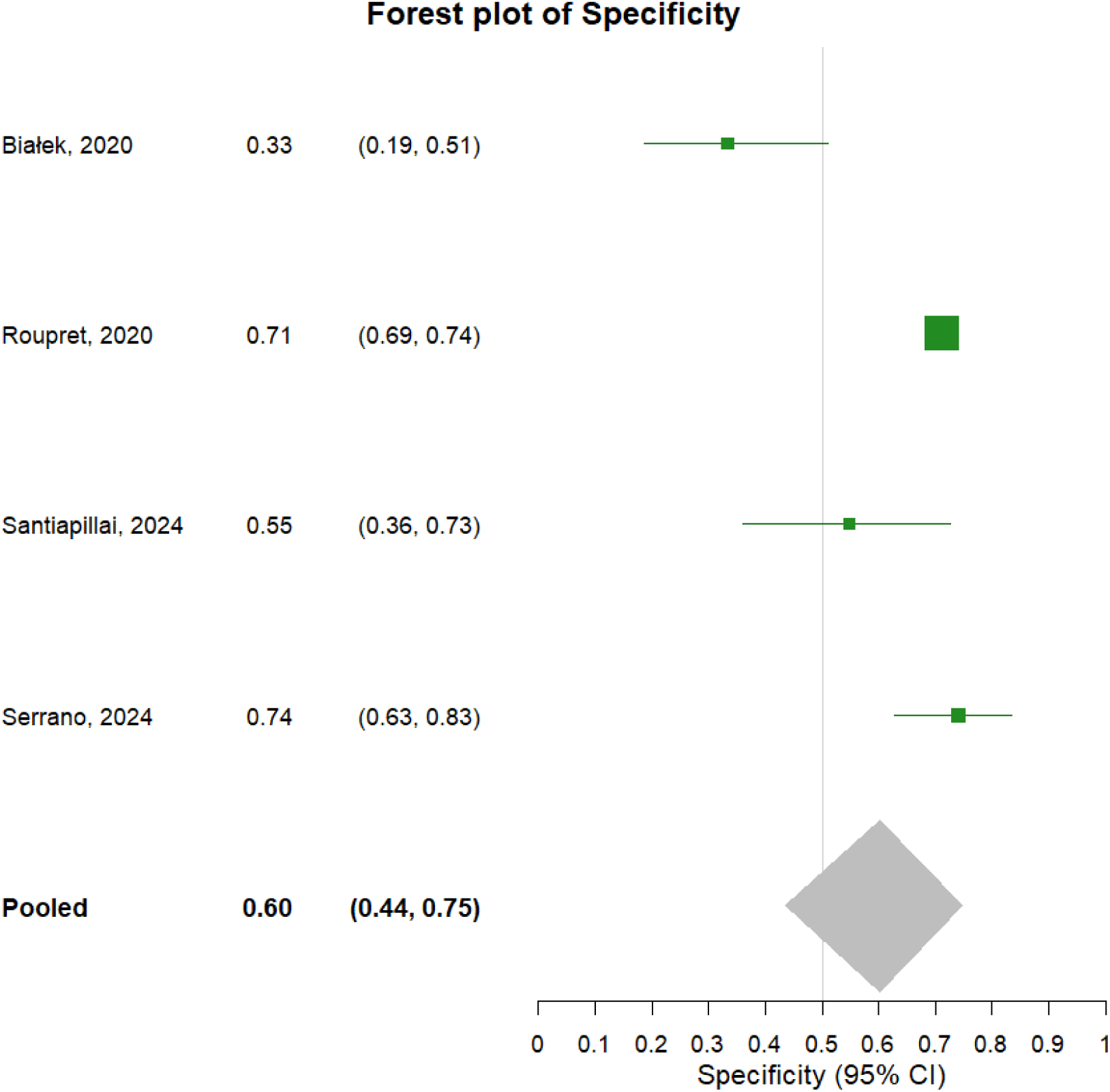
Forest plot of ADXbladder (specificity)

**Figure 9.**
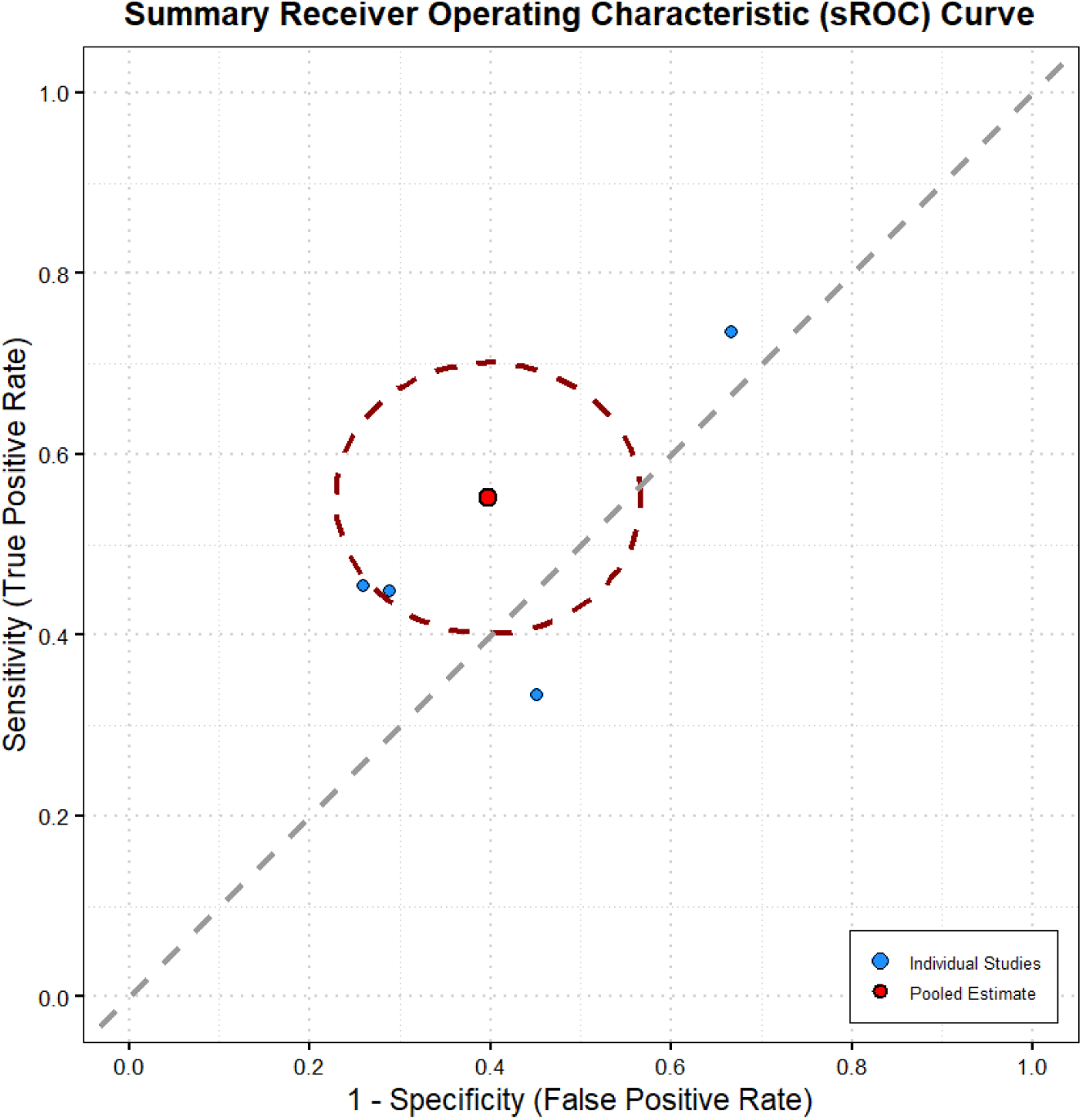
Summary sensitivity and specificity curve for ADXbladder

#### Diagnostic performance of Uromonitor

Six studies were used in the meta-analysis regarding the Uromonitor (Table 2). The pooled sensitivity and specificity estimates with confidence intervals of the six studies were 0.77 (0.61, 0.88) and 0.96 (0.91, 0.98), respectively (Figures 10 & 11). Sensitivity point estimates ranged from 0.49 to 0.93 and specificity point estimates ranged from 0.87 to 0.99 (Figure 12).

**Figure 10.**
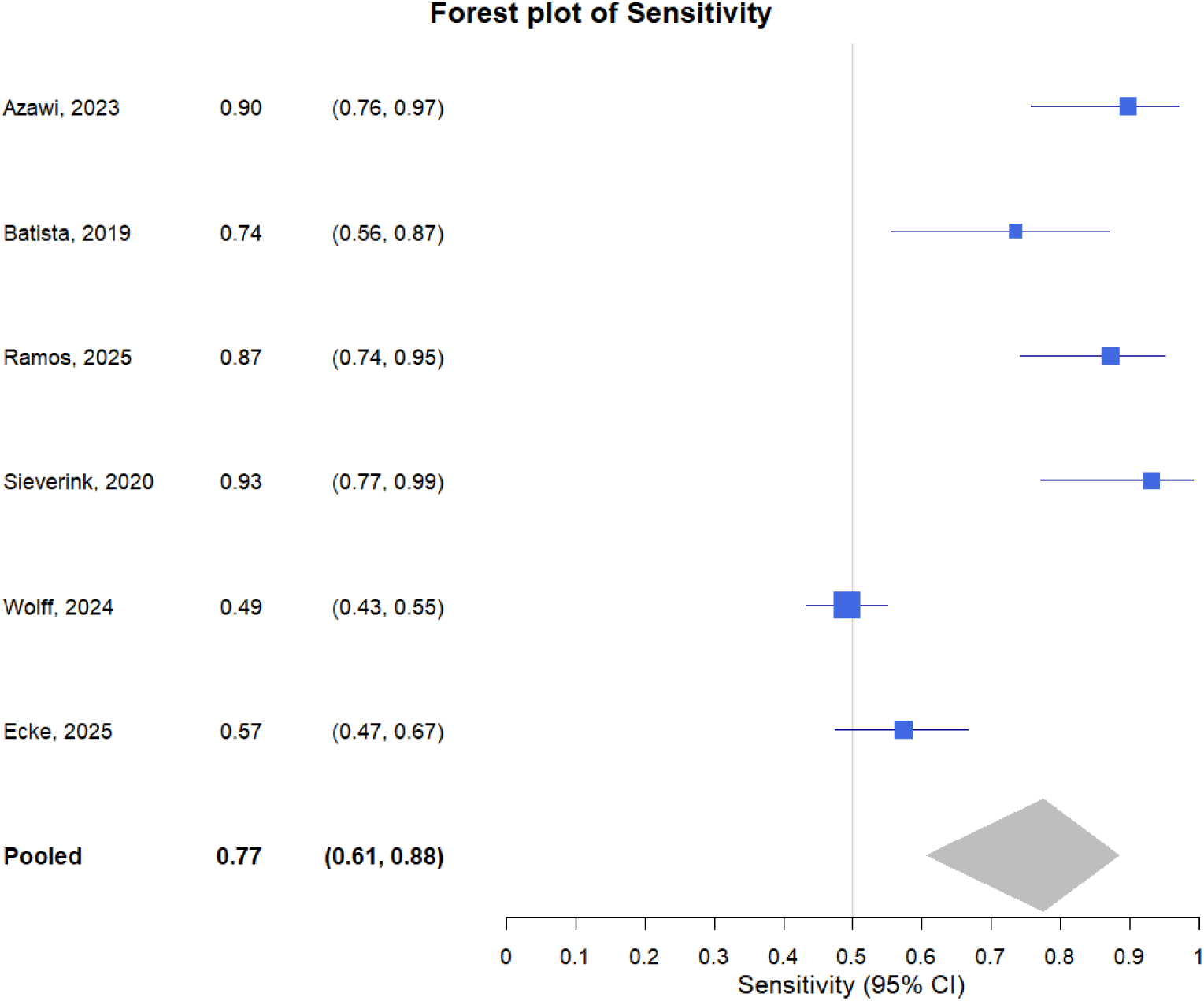
Forest plot of Uromonitor (sensitivity)

**Figure 11.**
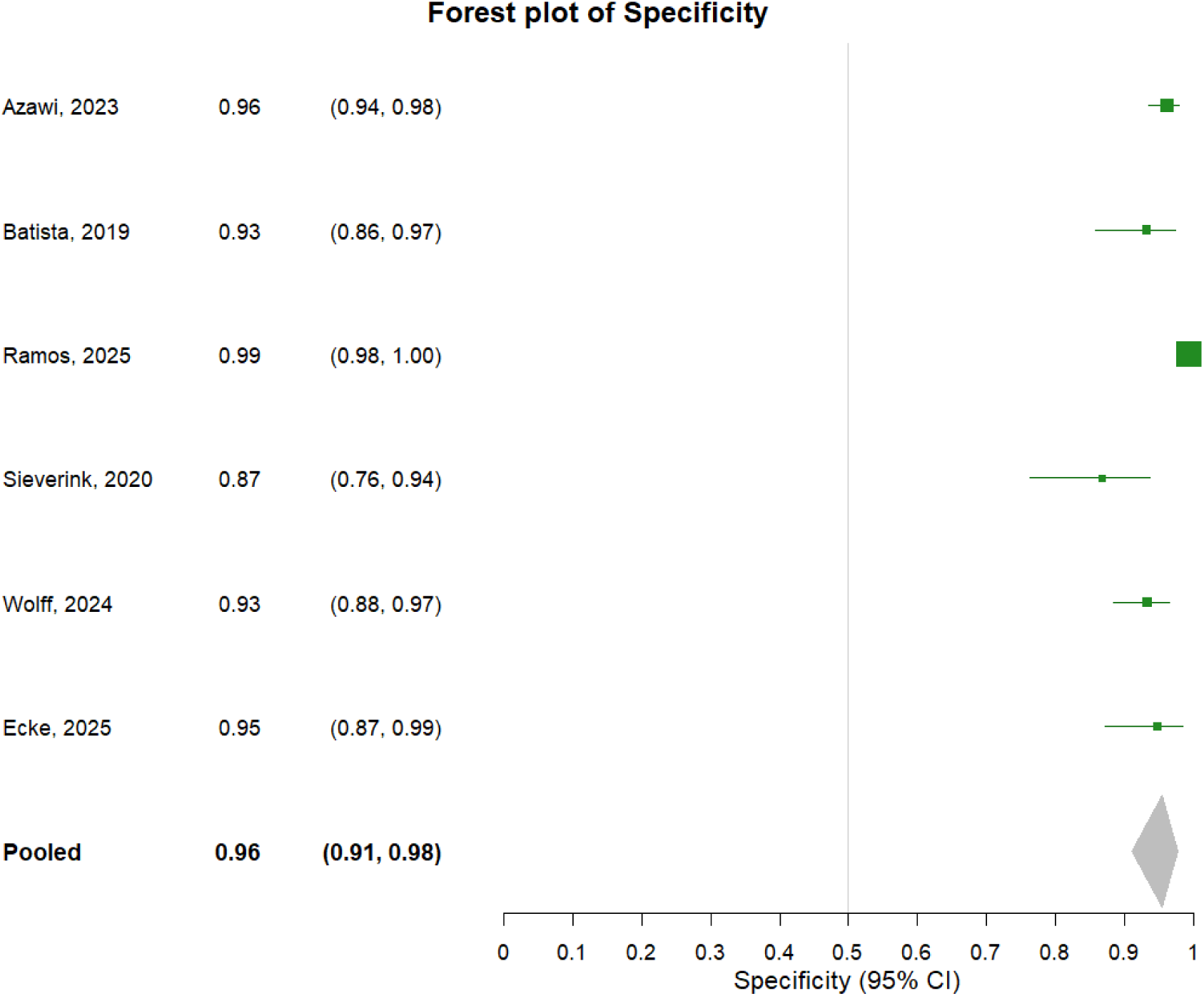
Forest plot of Uromonitor (specificity)

**Figure 12.**
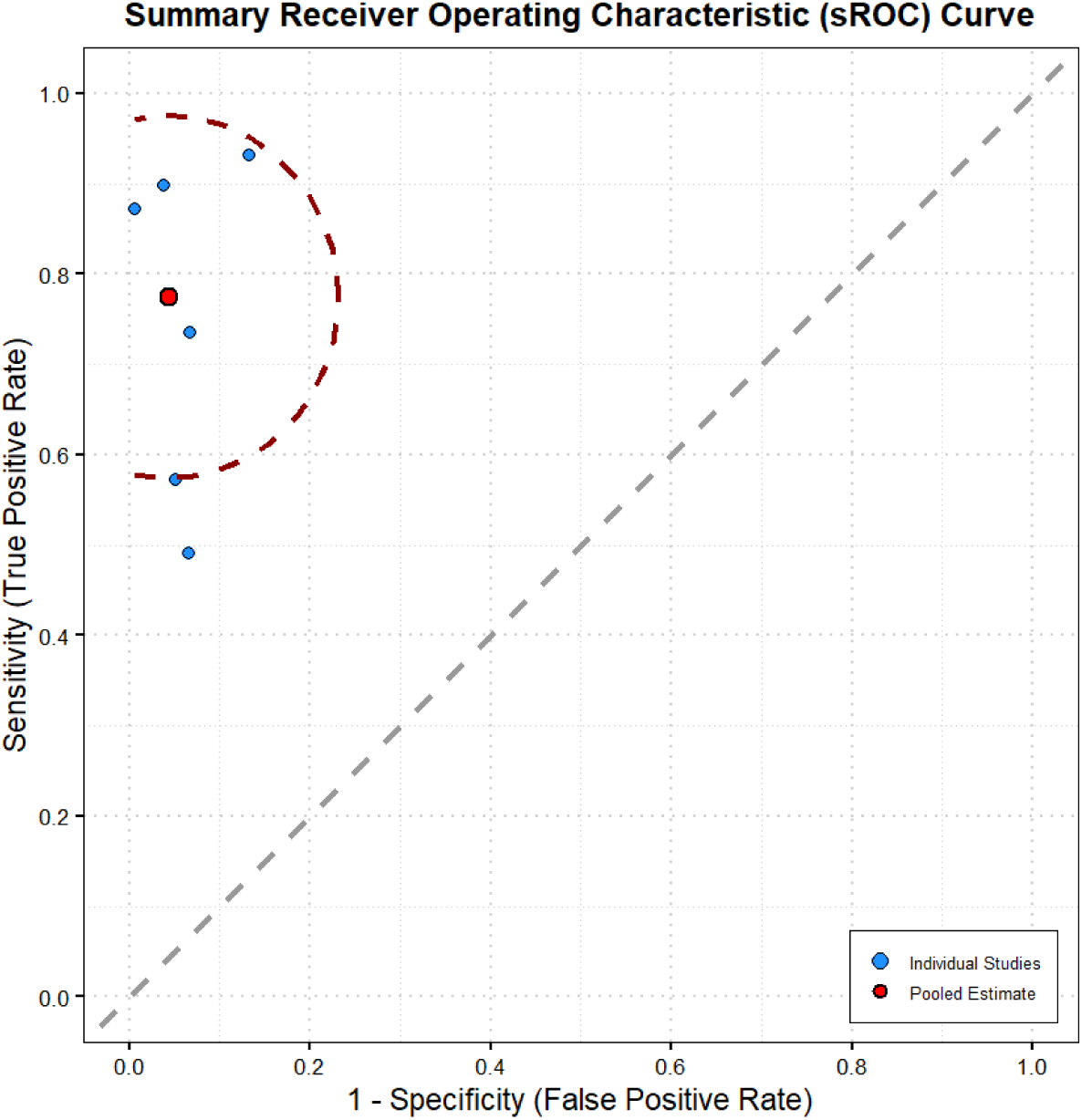
Summary sensitivity and specificity curve for Uromonitor

### Subgroup Analysis

#### High-Grade Tumors

The pooled sensitivity and specificity estimates with confidence intervals for high-grade tumors are as follows: Xpert Bladder Cancer, 0.84 (0.76, 0.90) and 0.75 (0.71, 0.79), respectively (Figures 13 & 14); Bladder EpiCheck, 0.88 (0.79, 0.93) and 0.93 (0.81, 0.97), respectively (Figures 15 & 16); ADXBladder, 0.78 (0.58, 0.90) and 0.51 (0.23, 0.78), respectively (Figures 17 & 18).

**Figure 13.**
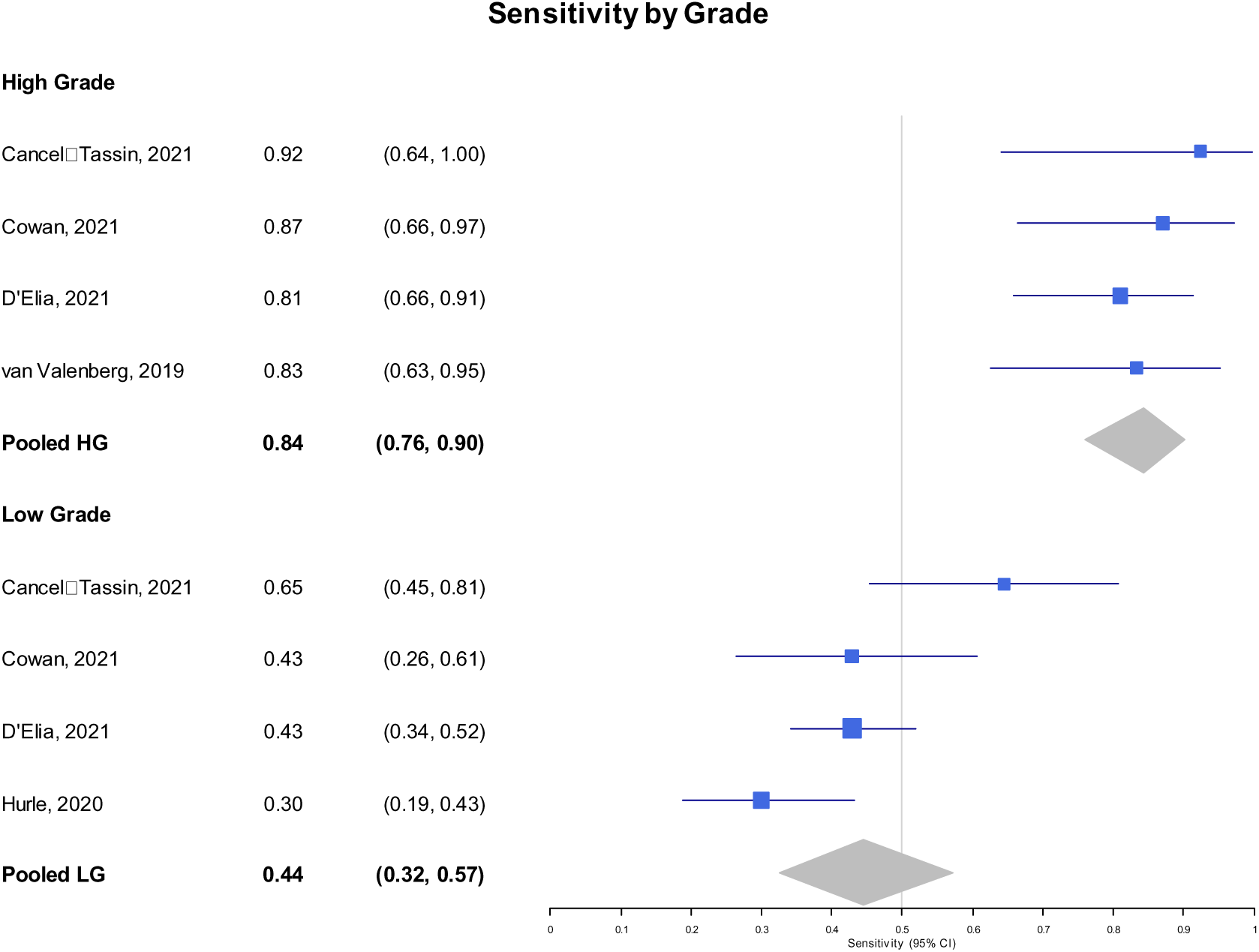
Forest plot of Xpert Bladder Cancer by Grade (sensitivity).

**Figure 14.**
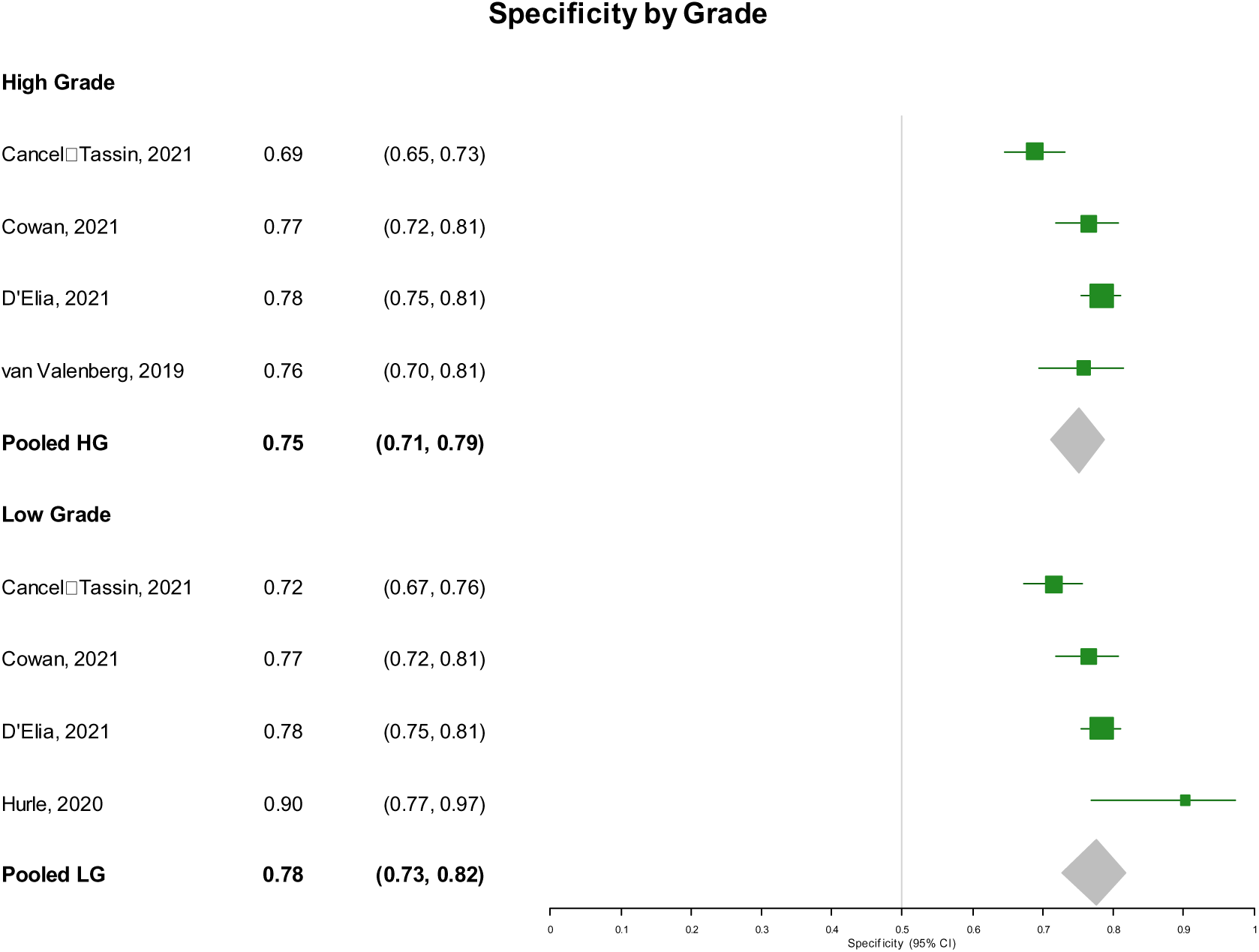
Forest plot of Xpert Bladder Cancer by Grade (specificity).

**Figure 15.**
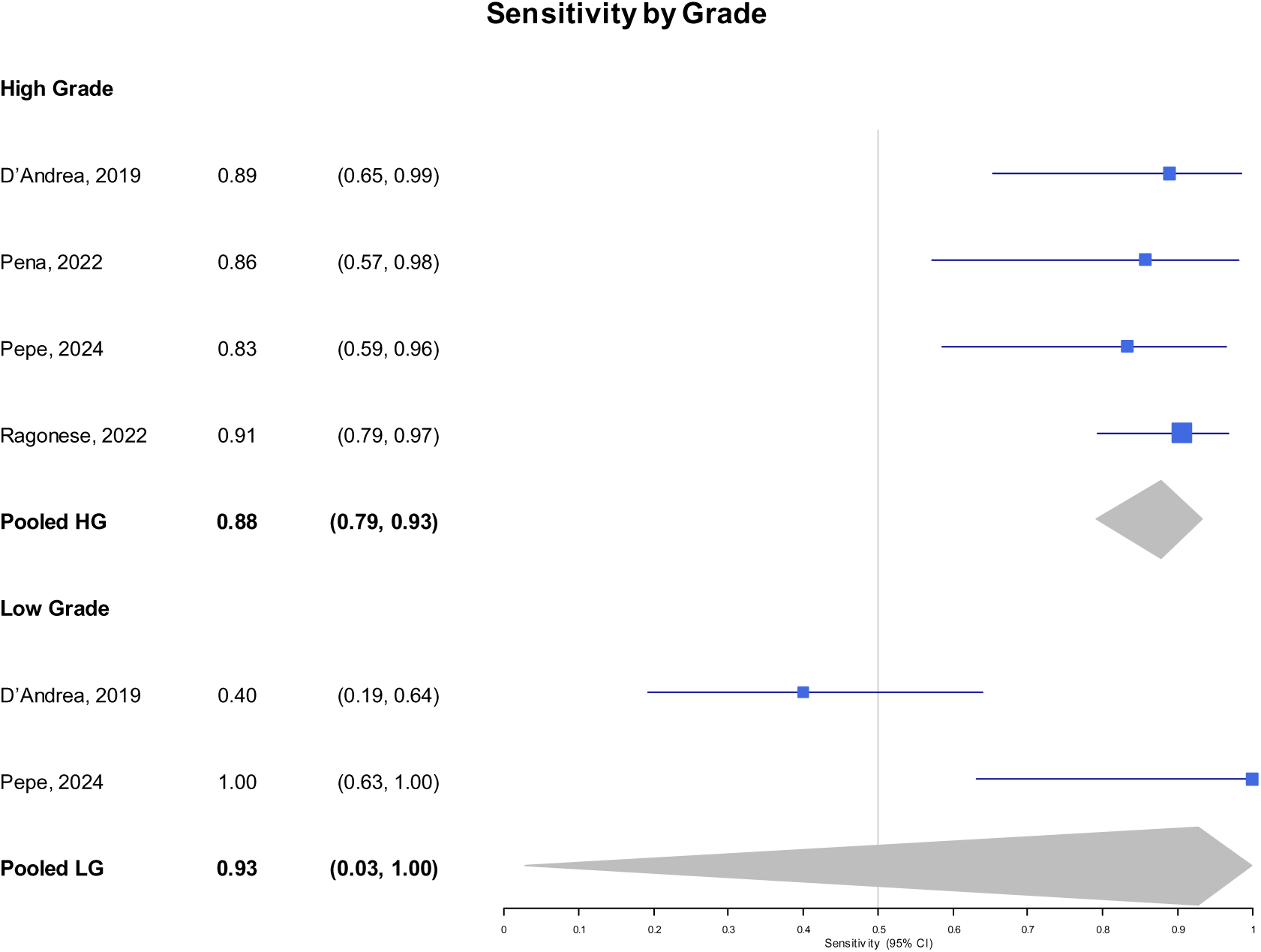
Forest plot of Bladder Epicheck by Grade (sensitivity).

**Figure 16.**
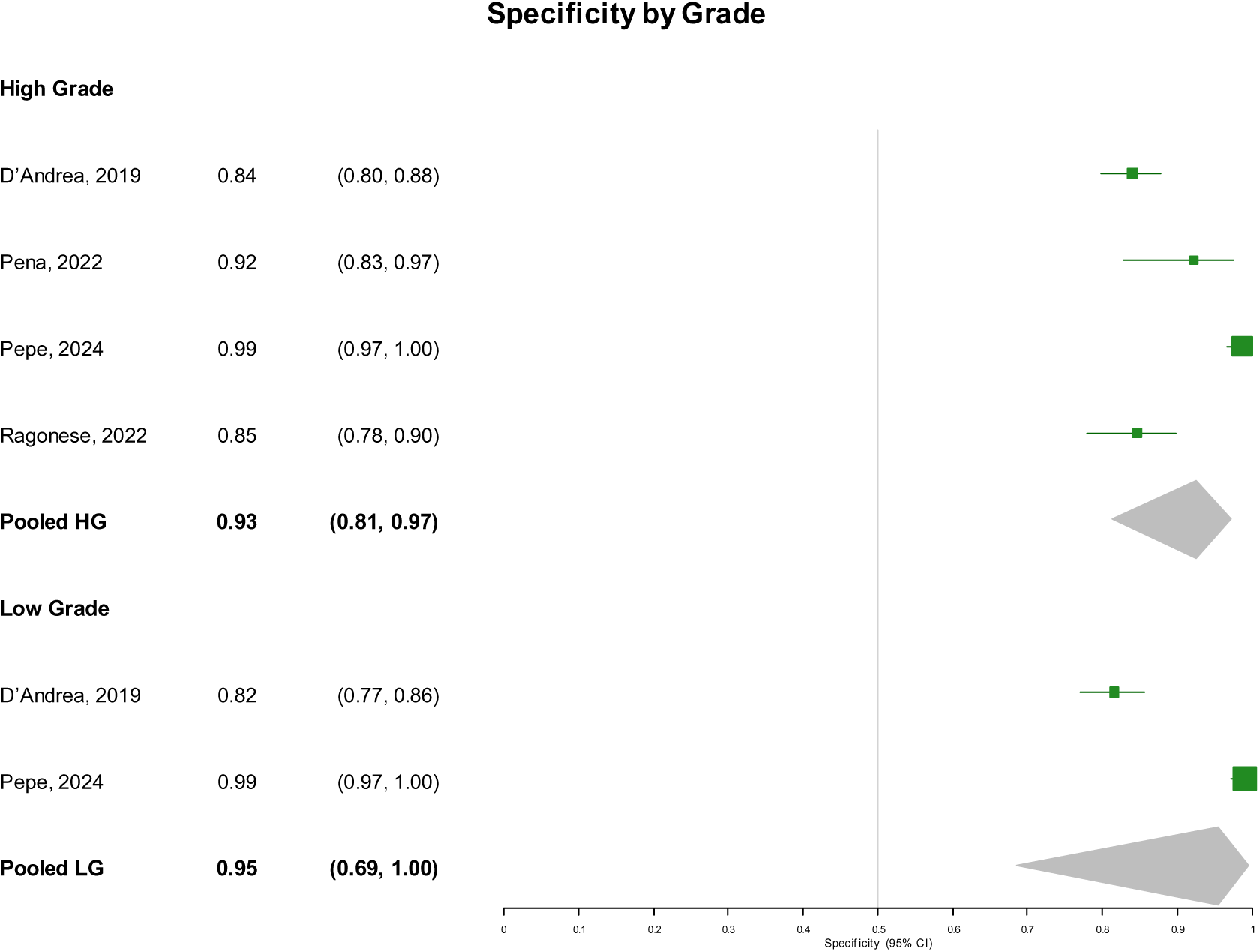
Forest plot of Bladder Epicheck by Grade (specificity).

**Figure 17.**
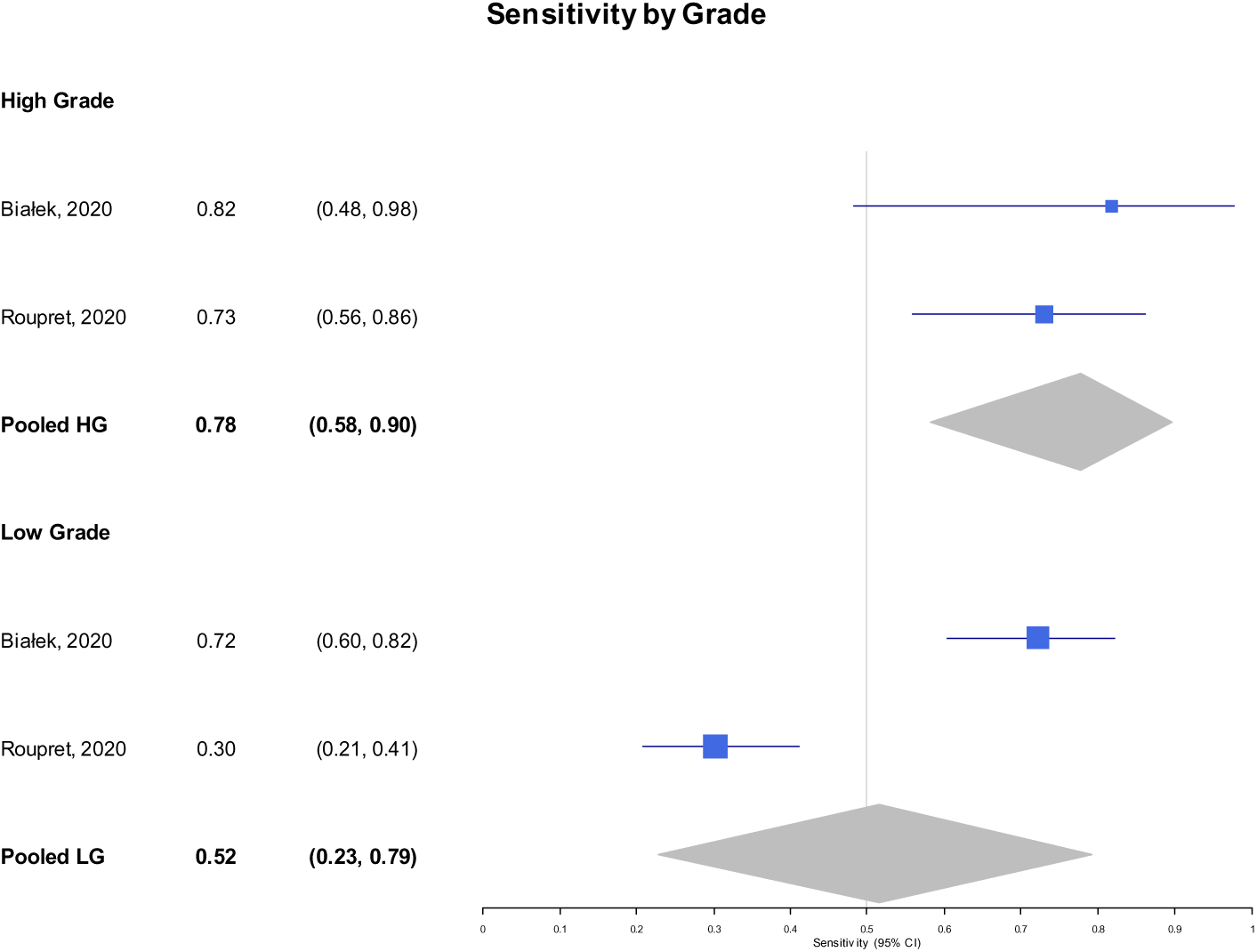
Forest plot of ADXbladder by Grade (sensitivity).

**Figure 18.**
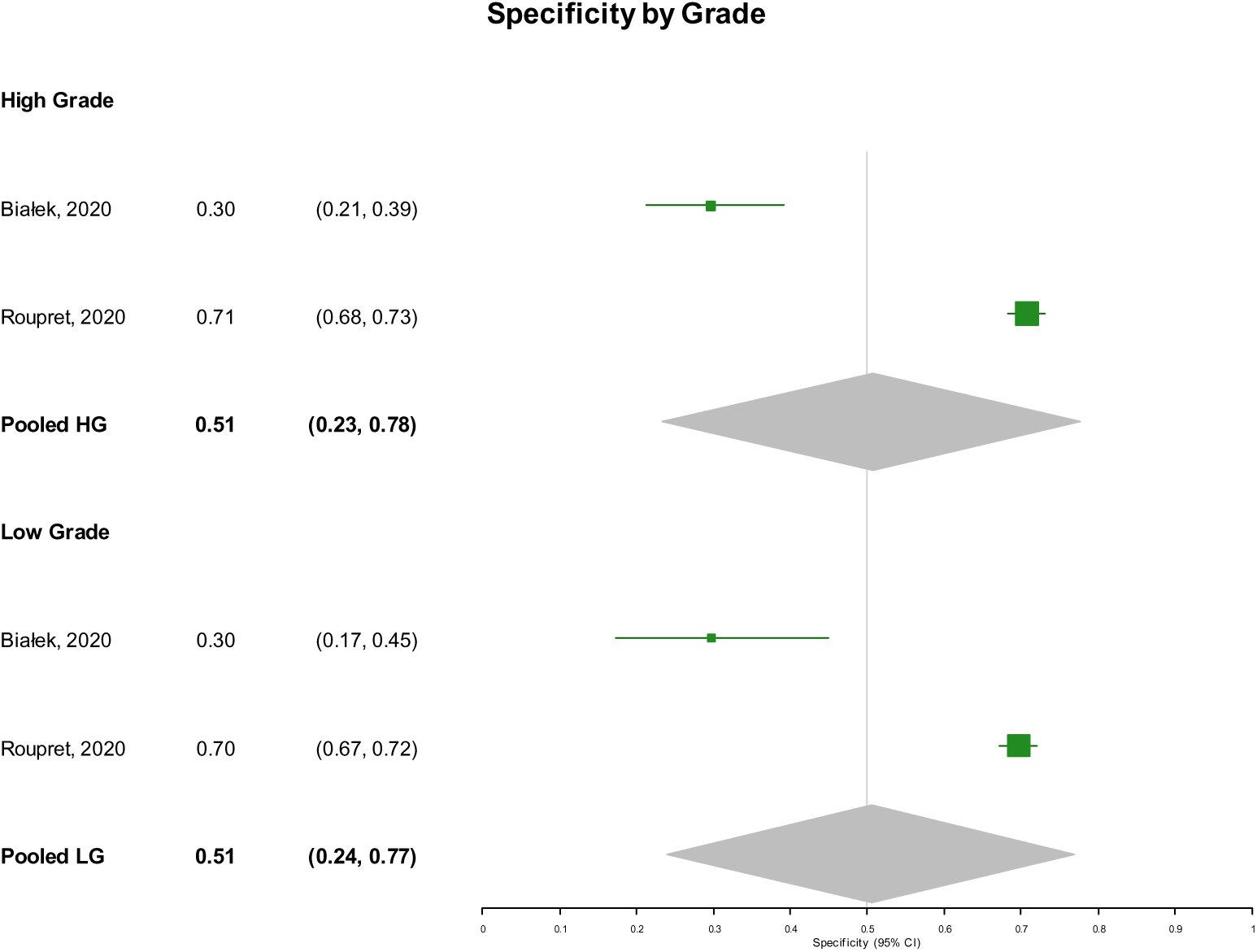
Forest plot of ADXbladder by Grade (specificity).

**Figure 19.** Forest plot of Uromonitor by Grade (sensitivity).

**Figure 20.** Forest plot of Uromonitor by Grade (specificity).

#### Low-Grade Tumors

The pooled sensitivity and specificity estimates with confidence intervals for low-grade tumors are as follows: Xpert Bladder Cancer, 0.44 (0.32, 0.57) and 0.78 (0.73, 0.82), respectively (Figures 13 & 14); Bladder EpiCheck, 0.93 (0.03, 1.00) and 0.95 (0.69, 1.00), respectively (Figures 15 & 16); ADXBladder, 0.52 (0.23, 0.79) and 0.51 (0.24, 0.77), respectively (Figures 17 & 18).

## Discussion

In the present meta-analysis we evaluated the diagnostic performance of five commercially-available UBTs in the diagnosis of BC recurrence (Xpert Bladder Cancer, Bladder Epicheck, ADXBladder, CxBladder Monitor, and Uromonitor). Thirty-six studies totally were met the inclusion criteria and Uromonitor, with three studies in its meta-analysis, exhibits the highest sensitivity and specificity, indicating the highest diagnostic performance among the remaining tests. In contrast, the ADXBladder using five studies appears to have the lowest sensitivity and a relatively low specificity. Bladder Epicheck and Xpert Bladder Cancer rank in an intermediate position, with moderate sensitivity and good specificity. These tests show satisfactory diagnostic performance, but their sensitivity may affect their clinical application. Uromonitor showed greater heterogeneity between studies, which may indicate different participant characteristics, such as different stages of cancer.

Although several UBTs have been developed for BC surveillance, limited comprehensive evaluations exits. Sharma et. al. [57], conducted a systematic review and meta-analysis assessing the diagnostic performance of Xpert BC in 2896 patients across 11 prospective NMIBC studies, reporting a sensitivity and specificity of 73% and 77%, respectively. Xpert BC showed better performance for high-grade tumors (sensitivity 0.86, specificity 0.78, AUC 0.87) than for low-grade tumors (sensitivity 0.58, specificity 0.79, AUC 0.79). While it may reduce cystoscopy frequency in high-grade disease, it cannot fully replace it. In addition, the diagnostic accuracy of Uromonitor was evaluated using four studies with 1190 urological examinations [58], reporting sensitivity, specificity, PPV, NPV of 80.2%, 96.9%, 82.1 % and 96.6 % respectively, and an AUC (95 % CI: 0.851-0.921), supporting its use in oncological follow-up of NMIBC patients. A broader synthesis of non-FDA-approved UBTs in 22 studies involving 7330 patients [59], reported pooled sensitivity up to 93% and pooled specificity up to 84%. However, these studies present some limitations included variability in LDR cut-off values, heterogeneity in patient inclusion, recurrence rates and tumour grade, as well as a lack of standardised cutoffs and underrepresentation of CIS cases. There were incomplete patient characterization, inability to perform subgroup analyses and lack of data for test reproducibility, oncologic outcomes and cost-effectiveness. Additionally, some studies did not report blinding procedures and a small total number of cases was included overall.

The limitations of the study lie in the small number of studies used to evaluate mainly Uromonitor with three studies and ADXBladder which included five studies. In addition, for CxBladder Monitor, there was only one study, which does not allow for the extraction of generalized conclusions. Finally, the differences between the studies, presenting heterogeneity, may affect the pooled estimate, especially in meta-analyses with a small number of individual studies. In particular, there is heterogeneity between studies, as most studies include patients specifically diagnosed with NMIBC, while in 6 studies the population suffers from bladder cancer without explicitly distinguishing the type of NMIBC [53], [55], [60], [61], [62], [63]. There are also two studies that examine a population with a history of urothelial carcinoma [50], [64]. Therefore, it is recommended to conduct a sensitivity analysis by population category.

## Conclusions

In conclusion, this study analysed the diagnostic performance of commercially available urine biomarker tests for bladder cancer recurrence surveillance, examining four meta-analyses for the Xpert Bladder Cancer, Bladder Epicheck, ADXBladder and Uromonitor tests. The results indicate that the Bladder Epicheck and Xpert Bladder Cancer tests have the highest performance compared to the others, and satisfactory diagnostic accuracy for the diagnosis of bladder cancer recurrence. However, all four tests can be used in combination with established diagnostic practices, such as cystoscopy, to help additionally improve patient monitoring.

Further research can be carried out with individual meta-analyses in a specific population of studies, i.e. in patients with high-or low-grade cancer, in order to evaluate the diagnostic ability of the tests in these cases as well. Sensitivity analysis is also suggested, in which studies that evaluate urothelial carcinoma in general will be excluded. Additional research is required to determine whether these tests can be effectively used, in combination with other diagnostic methods, to improve the detection of cancer recurrence. It is possible that the combination of biomarkers with imaging methods could improve the diagnosis of recurrence. Finally, the development of new biomarker technologies that will show both increased sensitivity and specificity may compete with the current main surveillance method (cystoscopy) in terms of diagnostic ability, but also degrade it hierarchically in the future.

## Data Availability

All data produced in the present study are available upon reasonable request to the authors

