## Supplementary material for "Diagnostic Test Accuracy of Commercially Available Tests for The Recurrence of Bladder Cancer: A Systematic Review and Meta-Analysis"

**Supplementary Table 1. PRISMA 2020 Checklist.**

| **Section and Topic** | **Item #** | **Checklist item** | **Location where item is reported** |
| --- | --- | --- | --- |
| **TITLE** | | |  |
| Title | 1 | Identify the report as a systematic review. |  |
| **ABSTRACT** | | |  |
| Abstract | 2 | See the PRISMA 2020 for Abstracts checklist. |  |
| **INTRODUCTION** | | |  |
| Rationale | 3 | Describe the rationale for the review in the context of existing knowledge. |  |
| Objectives | 4 | Provide an explicit statement of the objective(s) or question(s) the review addresses. |  |
| **METHODS** | | |  |
| Eligibility criteria | 5 | Specify the inclusion and exclusion criteria for the review and how studies were grouped for the syntheses. |  |
| Information sources | 6 | Specify all databases, registers, websites, organisations, reference lists and other sources searched or consulted to identify studies. Specify the date when each source was last searched or consulted. |  |
| Search strategy | 7 | Present the full search strategies for all databases, registers and websites, including any filters and limits used. |  |
| Selection process | 8 | Specify the methods used to decide whether a study met the inclusion criteria of the review, including how many reviewers screened each record and each report retrieved, whether they worked independently, and if applicable, details of automation tools used in the process. |  |
| Data collection process | 9 | Specify the methods used to collect data from reports, including how many reviewers collected data from each report, whether they worked independently, any processes for obtaining or confirming data from study investigators, and if applicable, details of automation tools used in the process. |  |
| Data items | 10a | List and define all outcomes for which data were sought. Specify whether all results that were compatible with each outcome domain in each study were sought (e.g. for all measures, time points, analyses), and if not, the methods used to decide which results to collect. |  |
|  | 10b | List and define all other variables for which data were sought (e.g. participant and intervention characteristics, funding sources). Describe any assumptions made about any missing or unclear information. |  |
| Study risk of bias assessment | 11 | Specify the methods used to assess risk of bias in the included studies, including details of the tool(s) used, how many reviewers assessed each study and whether they worked independently, and if applicable, details of automation tools used in the process. |  |
| Effect measures | 12 | Specify for each outcome the effect measure(s) (e.g. risk ratio, mean difference) used in the synthesis or presentation of results. |  |
| Synthesis methods | 13a | Describe the processes used to decide which studies were eligible for each synthesis (e.g. tabulating the study intervention characteristics and comparing against the planned groups for each synthesis (item #5)). |  |
|  | 13b | Describe any methods required to prepare the data for presentation or synthesis, such as handling of missing summary statistics, or data conversions. |  |
|  | 13c | Describe any methods used to tabulate or visually display results of individual studies and syntheses. |  |
|  | 13d | Describe any methods used to synthesize results and provide a rationale for the choice(s). If meta-analysis was performed, describe the model(s), method(s) to identify the presence and extent of statistical heterogeneity, and software package(s) used. |  |
|  | 13e | Describe any methods used to explore possible causes of heterogeneity among study results (e.g. subgroup analysis, meta-regression). |  |
|  | 13f | Describe any sensitivity analyses conducted to assess robustness of the synthesized results. |  |
| Reporting bias assessment | 14 | Describe any methods used to assess risk of bias due to missing results in a synthesis (arising from reporting biases). |  |
| Certainty assessment | 15 | Describe any methods used to assess certainty (or confidence) in the body of evidence for an outcome. |  |
| **RESULTS** | | |  |
| Study selection | 16a | Describe the results of the search and selection process, from the number of records identified in the search to the number of studies included in the review, ideally using a flow diagram. |  |
|  | 16b | Cite studies that might appear to meet the inclusion criteria, but which were excluded, and explain why they were excluded. |  |
| Study characteristics | 17 | Cite each included study and present its characteristics. |  |
| Risk of bias in studies | 18 | Present assessments of risk of bias for each included study. |  |
| Results of individual studies | 19 | For all outcomes, present, for each study: (a) summary statistics for each group (where appropriate) and (b) an effect estimate and its precision (e.g. confidence/credible interval), ideally using structured tables or plots. |  |
| Results of syntheses | 20a | For each synthesis, briefly summarise the characteristics and risk of bias among contributing studies. |  |
|  | 20b | Present results of all statistical syntheses conducted. If meta-analysis was done, present for each the summary estimate and its precision (e.g. confidence/credible interval) and measures of statistical heterogeneity. If comparing groups, describe the direction of the effect. |  |
|  | 20c | Present results of all investigations of possible causes of heterogeneity among study results. |  |
|  | 20d | Present results of all sensitivity analyses conducted to assess the robustness of the synthesized results. |  |
| Reporting biases | 21 | Present assessments of risk of bias due to missing results (arising from reporting biases) for each synthesis assessed. |  |
| Certainty of evidence | 22 | Present assessments of certainty (or confidence) in the body of evidence for each outcome assessed. |  |
| **DISCUSSION** | | |  |
| Discussion | 23a | Provide a general interpretation of the results in the context of other evidence. |  |
|  | 23b | Discuss any limitations of the evidence included in the review. |  |
|  | 23c | Discuss any limitations of the review processes used. |  |
|  | 23d | Discuss implications of the results for practice, policy, and future research. |  |
| **OTHER INFORMATION** | | |  |
| Registration and protocol | 24a | Provide registration information for the review, including register name and registration number, or state that the review was not registered. |  |
|  | 24b | Indicate where the review protocol can be accessed, or state that a protocol was not prepared. |  |
|  | 24c | Describe and explain any amendments to information provided at registration or in the protocol. |  |
| Support | 25 | Describe sources of financial or non-financial support for the review, and the role of the funders or sponsors in the review. |  |
| Competing interests | 26 | Declare any competing interests of review authors. |  |
| Availability of data, code and other materials | 27 | Report which of the following are publicly available and where they can be found: template data collection forms; data extracted from included studies; data used for all analyses; analytic code; any other materials used in the review. |  |

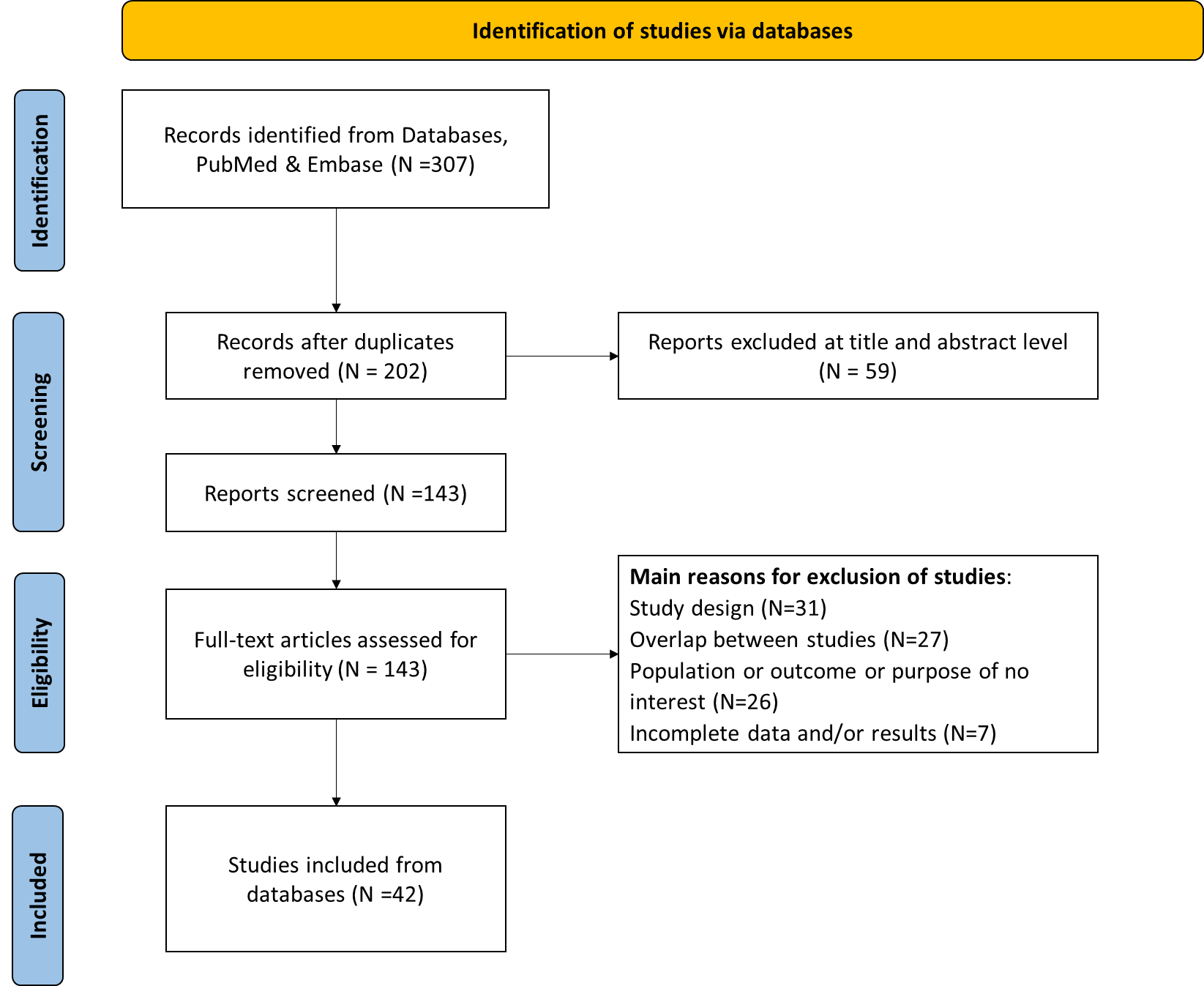

**Supplementary Figure 1. Flow chart of literature search and study selection according to PRISMA 2020 for all records together.**

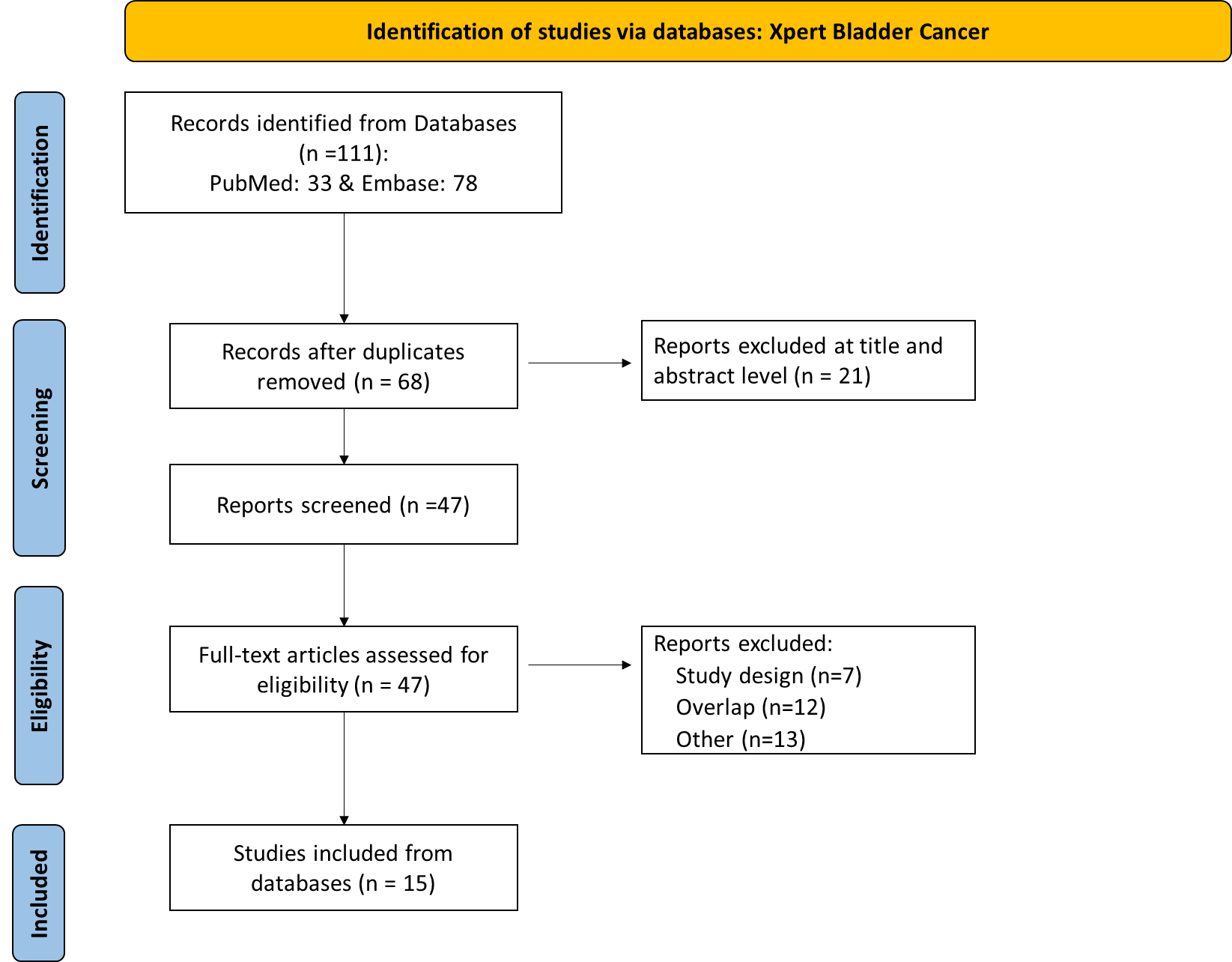

**Supplementary Figure 2. Flow chart of literature search and study selection according to PRISMA 2020 regarding the XPERT Bladder Cancer.**

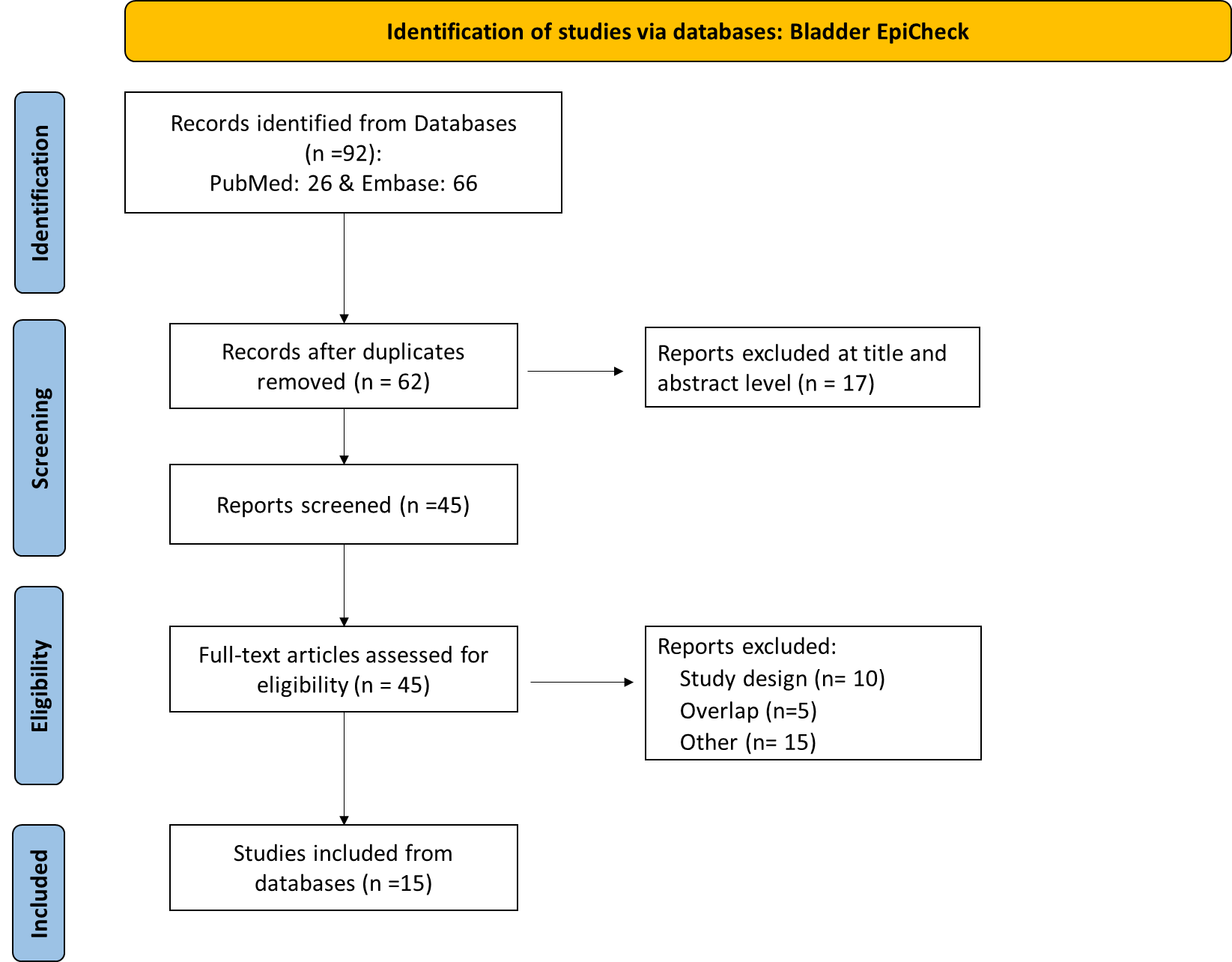

**Supplementary Figure 3. Flow chart of literature search and study selection according to PRISMA 2020 regarding the Bladder Epicheck**

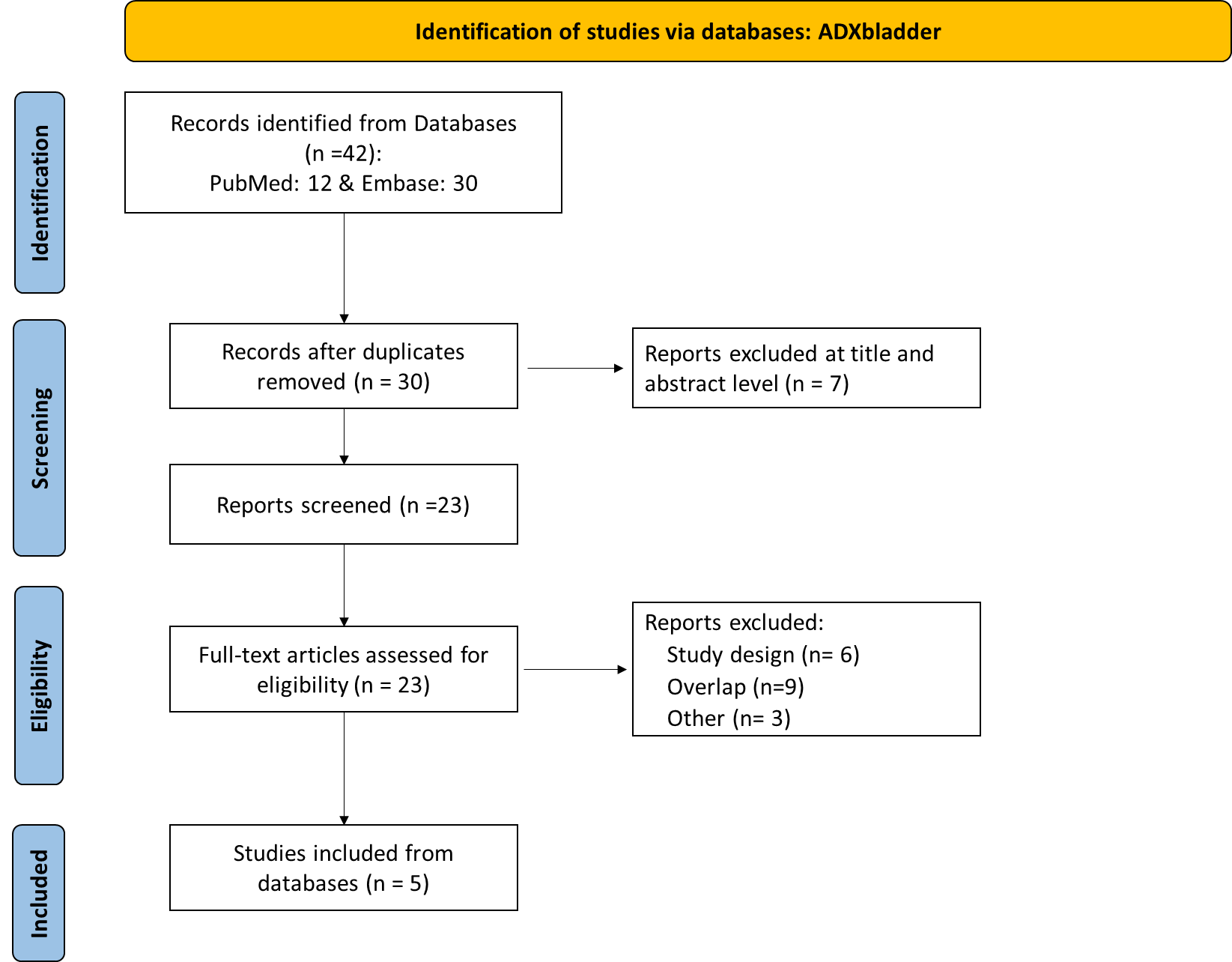

**Supplementary Figure 4. Flow chart of literature search and study selection according to PRISMA 2020 regarding the ADXbladder.**

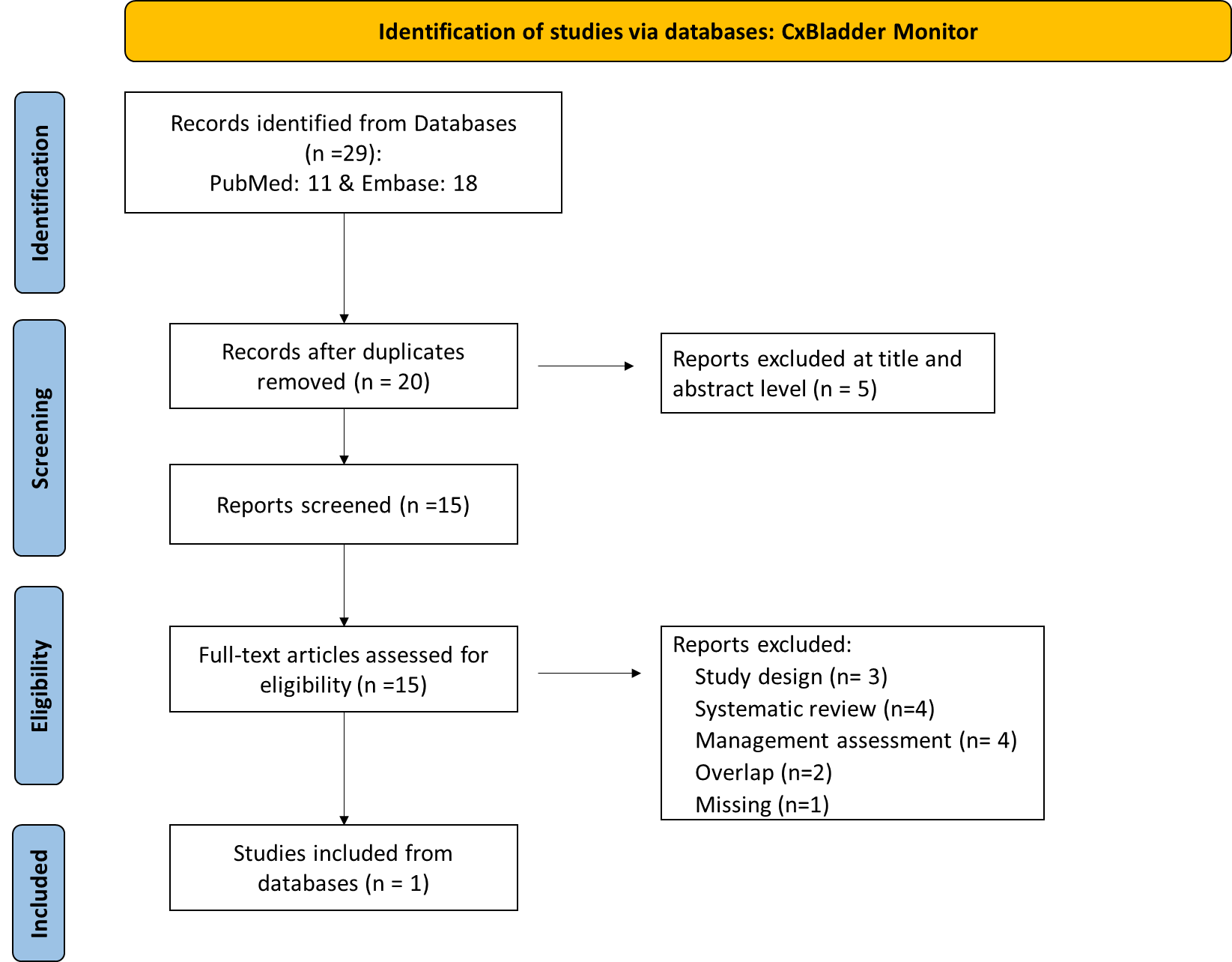

**Supplementary Figure 5. Flow chart of literature search and study selection according to PRISMA 2020 regarding the CxBladder Monitor.**

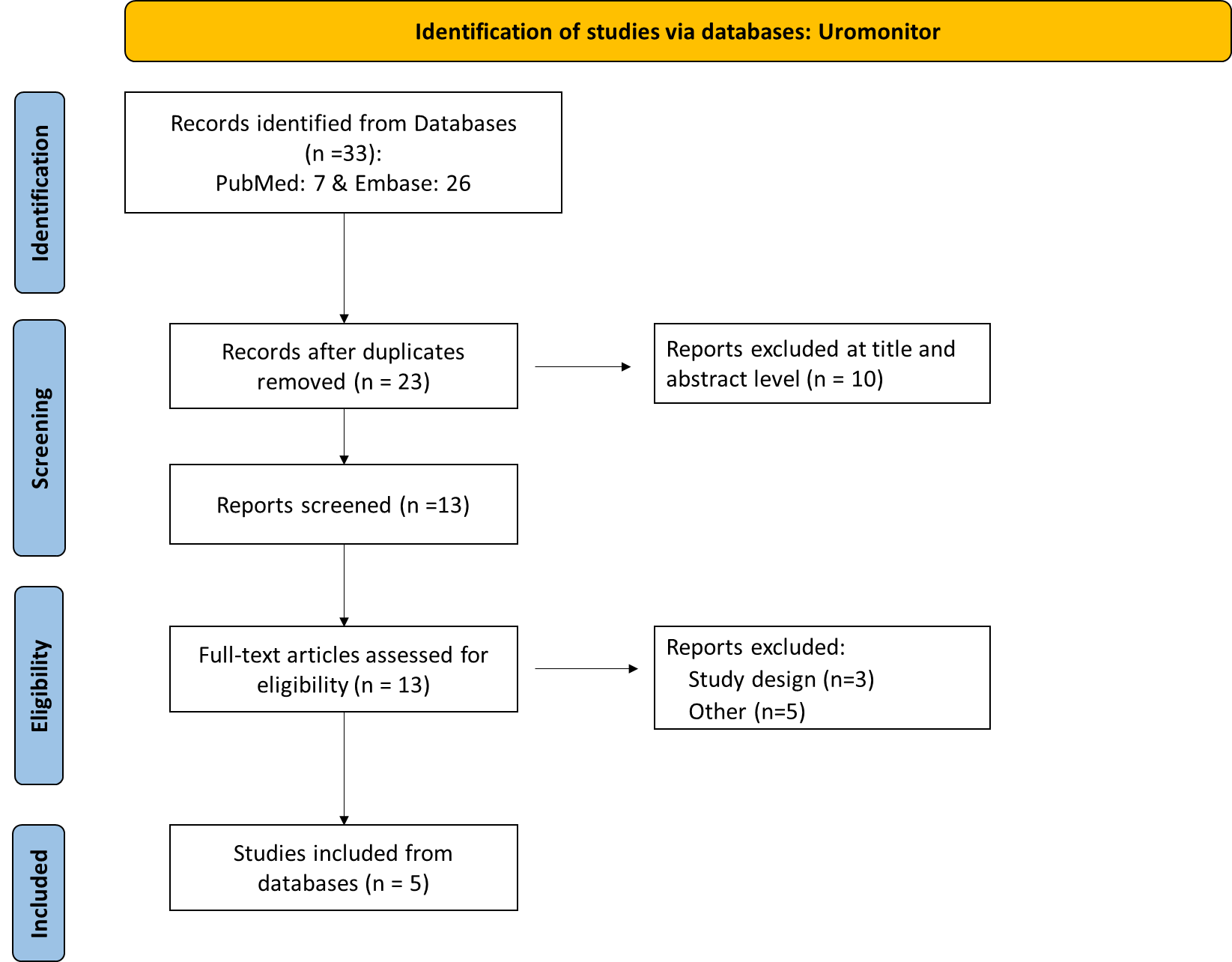

**Supplementary Figure 6. Flow chart of literature search and study selection according to PRISMA 2020 regarding the Uromonitor.**

**Supplementary Table 2. Reasons for exclusion of studies evaluated in terms of their content in each diagnostic test.**

| **First author, year** | **Reasons for exclusion of studies that were assessed overall in terms of their content** |
| --- | --- |
| **Uromonitor** | |
| Bras, 2023 [1] | Out of topic |
| Kravchuk, 2024 [2] | Review and meta-analysis |
| Leao, 2019 [3] | Inappropriate outcome and population (initial diagnosis) |
| Lee and Kim, 2020 [4] | Review |
| Schulz, 2022 [5] | Review |
| Wirtz, 2022 [6] | Incomplete data |
| Wirtz, 2024 [7] | Out of topic |
| EAU, 2024 [8] | Out of topic |
| **CxBladder** | |
| Charpentier, 2021 [9] | Review |
| Chu, 2021 [10] | Out of topic |
| Freifeld, 2018 [11] | Incomplete data regarding the diagnostic test |
| Jordaens, 2023 [12] | Review |
| Krabbe, 2016 [13] | Review |
| Koya, 2020 [14] | Non-diagnostic evaluation |
| Kavalieris, 2017 | Overlap with the study Lotan 2017 |
| Lotan, 2016 [15] | Overlap with the study Lotan 2017 |
| Li, 2022 [16] | Inappropriate population |
| Lough, 2018 [17] | Clinical utility |
| Mikhaylenko, 2019 [18] | Foreign language article |
| Soorojebally, 2023 [19] | Review |
| Tan, 2016 [20] | Case report and Inappropriate population |
| **ADXBladder** | |
| Benderska-Soder, 2024 [21] | Review |
| Dudderidge, 2018 [22] | Inappropriate outcome |
| Dudderidge, 2019 [23] | Inappropriate outcome |
| Dudderidge (abstract code: P10-8), 2021 [24] | Overlap with the study Montanari, 2020 |
| Dudderidge (abstract code: P10-10), 2021 [24] | Out of topic (diagnostic accuracy of recurrence is not assessed) |
| Gontero, 2020 [25] | Overlap with the study Roupret 2020 |
| Gontero, 2020 [26] | Overlap with the study Montanari 2021 |
| Gontero, 2020 [27] | Overlap with the study Gontero 2020 [25] |
| Gontero, 2021 [28] | Overlap with the study Dudderidge (abstract code: P10-8) 2021 |
| Lee, 2020 [4] | Review |
| Montanari, 2020 [29] | Overlap with the study Roupret, 2020 |
| Montanari, 2021 [30] | Λείπουν αποτελέσματα για την υποτροπή |
| Palou, 2020 [31] | Overlap with the study Roupret 2020 |
| Roupret, 2020 [32] | Overlap with the study Roupret 2020 |
| Roupret, 2021 [33] | Overlap with the study Montanari |
| Schulz, 2022 [5] | Review |
| Sharma, 2021 [34] | Review and meta-analysis |
| Soorojebally, 2023 [19] | Review |
| Wolfs, 2020 [35] | Review |
| **Xpert Bladder Cancer** | |
| Benderska-Soder, 2024 [21] | Review |
| Breyer, 2023 [36] | Inappropriate population |
| Cancel-Tassin, 2019 [37] | Overlap with the study Cancel-Tassin 2021 |
| Charpentier, 2021 [9] | Review |
| D'Elia, 2018 [38] | Overlap with the study D’Elia 2021 |
| D'Elia, 2018 [39] | Overlap with the study D’Elia 2021 |
| D'Elia, 2019 [40] | Overlap with the study D’Elia 2020 |
| D'Elia, 2020 [41] | Overlap with the study D’Elia 2021 |
| D'Elia, 2020 [42] | Overlap with the study D’Elia 2020 |
| D’Elia, 2022 [43] | Inappropriate population |
| D’Elia, 2023 [44] | Inappropriate population |
| Dreyer, 2023 [45] | Incomplete results for recurrence |
| Elsawy, 2020 [46] | Overlap with the study Elsawy 2020 |
| Elsawy, 2020 [47] | Inappropriate population |
| Fasulo, 2021 [48] | Usability study and overlay with Fasulo, 2022 |
| Fasulo, 2022 [49] | Utility study |
| Hefermehl, 2021 [50] | Overlap with the study Singer |
| Hurle, 2019 [51] | Overlap with the study Hurle 2020 |
| Kassem, 2020 [52] | Error in timing sequence (test after diagnosis) |
| Lazzeri, 2019 [53] | Overlap with the study Hurle 2020 |
| Lee, 2020 [4] | Review |
| Miyake, 2019 [54] | Inappropriate design (editorial comment) |
| Pycha, 2023 [55] | Inappropriate population |
| Schmitz-Drager, 2023 [56] | Inappropriate population |
| Schulz, 2022 [5] | Review |
| Sharma, 2022 [57] | Review and meta-analysis |
| Valenberg, 2017 [58] | Overlap with the study Valenberg 2021 |
| Valenberg, 2021 [59] | Inappropriate population |
| Zagorac, 2019 [60] | Use of a different diagnostic test (UroVysion FISH) |
| Zagorac, 2021 | Use of a different diagnostic test (UroVysion FISH) |
| **Bladder Epicheck** | |
| Afferi, 2020 [61] | Review |
| Bassi, 2020 |  |
| Batista, 2020 [62] | Review |
| Benderska-Soder, 2024 [21] | Review |
| Boissier, 2020 [63] | Inappropriate population |
| Breda, 2019 [64] | Inappropriate population |
| D'Elia, 2023 [65] | Inappropriate population |
| Di Gianfrancesco, 2019 [66] | Overlap with the study Bassi 2020 |
| Fiorentino, 2023 [67] | Review |
| Fleshner, 2024 [68] | Review |
| Gallioli, 2021 [69] | Inappropriate population |
| Heard, 2024 [70] | Review |
| Kałuzewski, 2021 [71] | Non-diagnostic study – Inappropriate target |
| Lee, 2020 [4] | Review |
| Mancini, 2020 [72] | Review |
| Palermo, 2023 [73] | Inappropriate population |
| Pierconti, 2020 |  |
| Pierconti, 2021 [74] | Inappropriate population |
| Pierconti, 2021[75] | Incomplete data– Overlap with the study Pierconti 2020 |
| Pierconti, 2022 [76] | Overlap with the study Pierconti 2020 |
| Pierconti, 2022 [77] | Overlap with the study Pierconti 2020 |
| Pierconti, 2023 [78] | Incomplete data |
| Pycha, 2023 [55] | Inappropriate population |
| Racioppi, 2020 [79] | Overlap with the study Bassi 2020 |
| Ragonese, 2021 |  |
| Ragonese, 2020 [80] | Overlap with the study Ragonese 2022 |
| Ramon, 2018 [81] | Inappropriate population |
| Righetto, 2022 |  |
| Territo, 2022 [82] | Inappropriate population |
| Wasserstrom, 2016 |  |
| Witjes, 2018 [83] | Overlap with the study Witjes 2019 |
| Witjes, 2018 [84] | Overlap with the study Witjes 2018 |
| Witjes, 2019 [85] | Incomplete results for recurrence |
| Witjes, 2021 [86] | Out of topic |
| Wolfs, 2021 [35] | Review |

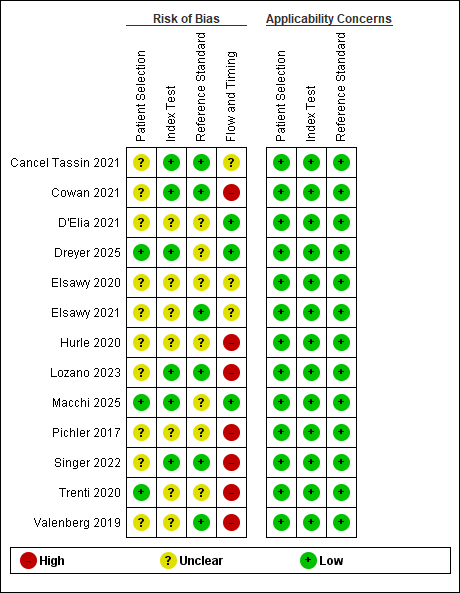

**Supplementary Figure 7. Risk of bias and applicability concerns summary: review authors' judgements about each domain for each included study for Xpert Bladder Cancer.**

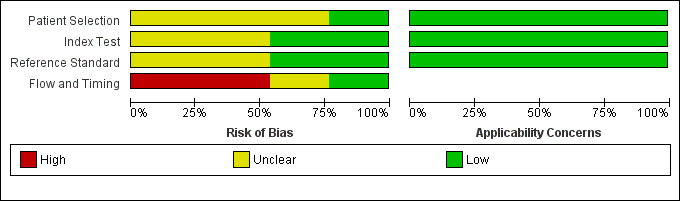

**Supplementary Figure 8. Risk of bias and applicability concerns graph: review authors' judgements about each domain presented as percentages across included studies for Xpert Bladder Cancer.**

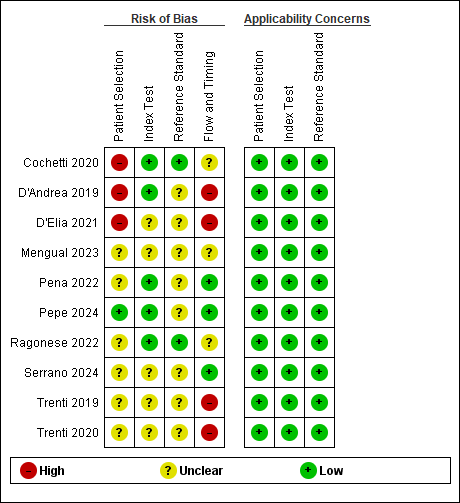

**Supplementary Figure 9. Risk of bias and applicability concerns summary: review authors' judgements about each domain for each included study for Bladder Epicheck.**

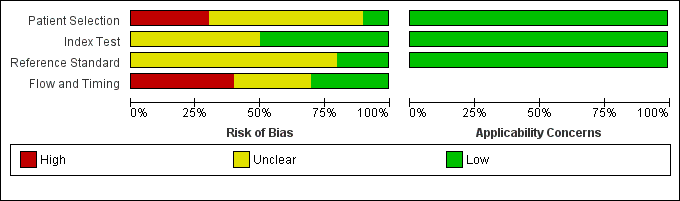

**Supplementary Figure 10. Risk of bias and applicability concerns graph: review authors' judgements about each domain presented as percentages across included studies for Bladder Epicheck.**

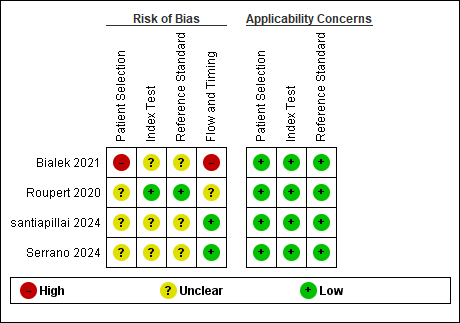

**Supplementary Figure 11. Risk of bias and applicability concerns summary: review authors' judgements about each domain for each included study for ADXBladder.**

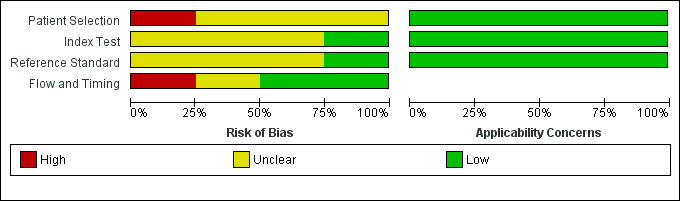

**Supplementary Figure 12. Risk of bias and applicability concerns graph: review authors' judgements about each domain presented as percentages across included studies for ADXBladder.**

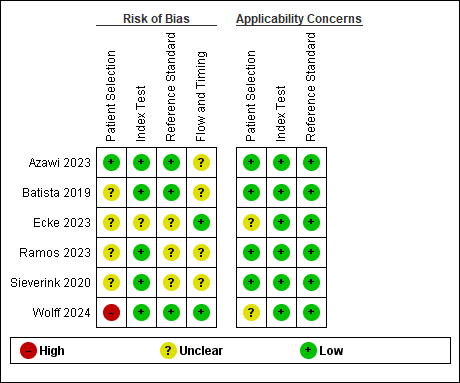

**Supplementary Figure 13. Risk of bias and applicability concerns summary: review authors' judgements about each domain for each included study for Uromonitor.**

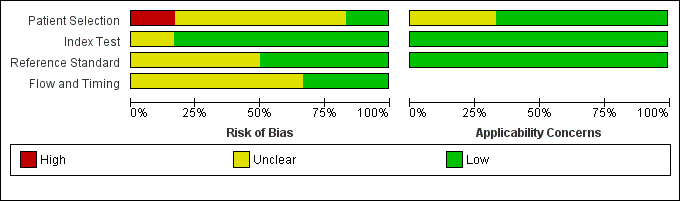

**Supplementary Figure 14. Risk of bias and applicability concerns graph: review authors' judgements about each domain presented as percentages across included studies for Uromonitor.**
